# National Longitudinal Mediators of Psychological Distress During Stringent COVID-19 Lockdown

**DOI:** 10.1101/2020.09.15.20194829

**Authors:** Joseph A. Bulbulia, Sofia D. Piven, Fiona Kate Barlow, Don E. Davis, Lara M. Greaves, Benjamin Highland, Carla A. Houkamau, Taciano L. Milfont, Danny Osborne, Nickola Christine Overall, John H. Shaver, Geoffrey Troughton, Marc Wilson, Kumar Yogeeswaran, Chris G. Sibley

## Abstract

We leverage powerful time-series data from a national longitudinal sample measured before the COVID-19 pandemic and during the world’s eighth most stringent COVID-19 lockdown (New Zealand, March-April 2020, *N* = 940) and apply Bayesian multilevel mediation models to rigorously test five theories of pandemic distress. Findings: (1) during lockdown, rest diminished distress; without rest psychological distress would have been ~ 1.74 times greater; (2) an elevated sense of community reduced distress, a little, but elevated government satisfaction was inert. Thus, the psychological benefits of lockdown extended to political discontents; (3) most lockdown distress arose from dissatisfaction from personal relationships. Social captivity, more than isolation, proved challenging; (4-5) Health and business satisfaction were stable; were they challenged substantially more distress would have ensued. Thus, lockdown benefited psychological health by affording safety, yet only because income remained secure. These national longitudinal findings clarify the mental health effects of stringent infectious disease containment.

---

Studies from Australia, China, Italy, Japan, and the United States find substantially elevated psychological distress during the early COVID-19 pandemic (Biddle et al., 2020; McGinty et al., 2020; Moccia et al., 2020; Qiu et al., 2020; Twenge & Joiner, 2020; Yamamoto et al., 2020). These studies propose three mechanisms to explain pandemic distress.

m1 Distress elevation from SARS-CoV-2 health risks (Qiu et al., 2020; Twenge & Joiner, 2020);
m2 Distress elevation from economic downturn (Twenge & Joiner, 2020);
m3 Distress elevation from social isolation (Van Bavel et al., 2020).

Early into the pandemic, New Zealanders also reported increased psychological distress, although at much lower levels (Sibley, Greaves, et al., 2020). Such a muted distress response is surprising because, at the time, New Zealanders faced health and economic uncertainties resembling those in countries that experienced severe distress (Appendix C). Additionally, New Zealand’s March/April lockdown was independently rated as the world’s 8*^th^* most stringent. It required sheltering-in-place with domestic cohabitants, and resulted in a sudden loss of ordinary business and social routines (Appendix C). Importantly, the effectiveness, duration, and economic consequences of New Zealand’s “go-hard, go-early” pandemic lockdown were unknown at the time, and presently remain unknown.

In their study of social attitudes and well-being during New Zealand’s early COVID-19 lockdown, Sibley, Greaves, et al. (2020) propose two mechanisms for mitigating pandemic distress:

m4 Distress buffering from greater institutional trust;
m5 Distress buffering from an elevated sense of civic community.

This proposal was based on the observation that institutional trust and civic attitudes were elevated during New Zealand’s early COVID-19 lockdown, as well as orthogonal evidence that positive public and civic attitudes affect mental health (Sibley, Greaves, et al., 2020).

Presently, a systematic quantitative understanding of psychological distress production and mitigation at the outset of a pandemic – one that uses longitudinal data within participants from pre-pandemic baselines to infer mechanisms of distress elevation and buffering – remains elusive. However, to the extent that mental health forms part of a government’s public health strategy, such a causal understanding is important wherever lethal infectious disease containment improves from repeated or prolonged lockdowns (Van Bavel et al., 2020).

Here, we leverage longitudinal data from a national probability sample of New Zealanders who responded to the New Zealand Attitudes and Values Study (NZAVS) during the first 18 days of New Zealand’s stringent COVID-19 lockdown from 26*^th^* March – 12*^th^* April 2020. We systematically quantify changes in psychological distress within the same individuals using their previous year’s responses as baseline. This approach affords a powerful tool for inference because:

1. The NZAVS has multiple indicators for parameters theorised to affect pandemic distress in five domains of current interest: **(m1) personal health**: health satisfaction, subjective health, rumination, and fatigue; **(m2) economic concerns**: satisfaction with the economy, standard of living, future security, and business satisfaction; **(m3) social relationships**: social support, social belonging, and satisfaction with personal relationships; **(m4) institutional trust**: satisfaction with the government, trust in politicians, willingness to engage with the police, and trust in the police; **(m5) civic attitudes**: satisfaction with social conditions, national identification, patriotism, and sense of neighbourhood community;
2. To measure psychological distress, the NZAVS uses the Kessler-6 scale, a diagnostically reliable measure used in clinical screening of anxiety and depression in many countries, including New Zealand (Kessler et al., 2002; Kessler et al., 2010; Krynen et al., 2013; Prochaska et al., 2012);
3. The NZAVS lockdown sample is demographically diverse and exhibits varied responses across the spectrum of theoretically relevant parameters.
4. Longitudinal mediation models quantify the extent to which changes in psychological distress were caused by changes in the availability and abundance of theoretically postulated pandemic distress parameters within the same individuals pre/during the pandemic lockdown. This affords systematic tests for many theories of pandemic distress. For example, if elevated health concerns were responsible for greater lockdown distress, as has been theorised, we would expect those who became more concerned about their health to present greater distress during lockdown compared with the previous year. If relationship dissatisfaction were to cause distress in the absence of economic and health concerns, we would infer that it was the effect of sheltering-in-place on relationship satisfaction that caused psychological distress, rather than the effects of health and economic uncertainties on relationship satisfaction that caused the psychological distress. (Indeed, this is what we observe).
5. Longitudinal mediation models quantitatively clarify theoretically important counterfactuals about the magnitudes of distress people would have experienced had distress-related parameters changed during lockdown. For example, although we observe that social belonging and satisfaction with future security were not elevated during lockdown, our statistical models indicate how much extra distress would have been caused had these parameters been compromised. Such clarity is important for policy-making during pandemics in the same way that surveying unknown waters is important for navigating ships: to avoid hazards we need to chart them.

## Method

### Sample

The New Zealand Attitudes and Values Study (NZAVS) is a national panel study that has been tracking social attitudes, personality, and health outcomes within the same individuals since 2009 (*Ns* = 6,441-47,951). Here, we conduct a novel analysis of the rolling NZAVS 2020 data-frame first used by Sibley, Greaves, et al. (2020) (pre-registration of that study: Sibley, Greaves, et al., 2020). The pre/post pandemic onset panel is composed of 940 participants who responded to both Wave 10 (2018) and Wave 11 (2020) of the NZAVS, which were taken respectively before and during the first eighteen days of New Zealand’s first COVID-19 lockdown (March 26*^th^* to April 12*^th^*). The Time 10 (2018) NZAVS contained responses from 47,951 participants (18,010 retained from one or more previous waves). A full comparison of the lockdown sample and full NZAVS pre-COVID-19 baseline reveals that the lockdown panel was similar to the full NZAVS national probability sample, showing good demographic coverage of the country’s population (see: Appendix E). We report sample descriptive statistics in D1.

### Mediators of Pandemic Distress

The five areas encompassing the indicators we used to investigate mediation (or “indirect”) effects on psychological distress are: (m1) personal health, (m2) economic concerns, (m3) social relationships, (m4) institutional trust, and (m5) civic attitudes. All indicators were obtained from Sibley, Greaves, et al. (2020)’s preregistered study describing overall changes in attitudes and well-being during New Zealand’s March/April 2020 lockdown: https://osf.io/e765a/. We used only those indicators identified in the preregistered study for which there was longitudinal information (the preregistered study took a propensity-score matching approach, and some indicators used in that study were not available in the previous NZAVS wave). We measured psychological distress using the Kessler-6 scale (Kessler et al., 2002), which exhibits strong diagnostic concordance for moderate and severe psychological distress (Kessler et al., 2010; Prochaska et al., 2012). We present full sample descriptive statistics for these indicators in Appendix D.

### Statistical models

We estimated multilevel mediation models using the bmlm package in R (Vuorre, 2017). Using standard mediation model notation:

1. a is the regression slope of the lockdown on a mediator (X→M)
2. b is the regression slope of the mediator on distress (M→Y)
3. c′ is the regression slope of the lockdown on distress (X→Y, the direct effect)
4. (me) is the mediated effect 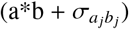
5. c is the total effect of X on Y (c= c′+ me).

In the mediation graphs, we report coefficients for the population means, with 95% Bayesian credible intervals in adjacent brackets. We also report coefficients for individual-level variation for all paths 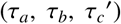 (see: Tables G1 – G19), these are indicated by “SD” coefficients on the directed graphs in Figures F1 – F19. Though reporting individual-level variation in not common in population health studies, identifying patterns of psychological distress in sub-populations is relevant to public mental health because there may be numerically many instances of mental health challenge at the margins of a population, which are not evident in the expected population means. Consider an analogy to infectious disease. The SARS-CoV2 virus causes mortality in less than 1% of infectious cases; it is lethal only at the population margin, yet capable of overwhelming a country’s health resources. Though less apparent than a piling up of corpses in hospital corridors, strong challenges to mental health among minority segments of a population during and following a pandemic must be documented and explained (on the 1918 influenza see: Menninger, 1920; Supplement A discusses individual differences). For a detailed description of our statistical models see Appendix D. The R scripts for this study are available at https://osf.io/5gza8/.

## Results

### Health mediators of lockdown distress

We do not find a reliable effect of the lockdown on health satisfaction (a = −0.07, [-0.17, 0.04], τ*_a_* = 0.15, [0.01, 0.42]), and so there was not a mediation effect of the lockdown on distress through health satisfaction (me = 0.01,[-0.03, 0.04]). However, we find health satisfaction reliably predicted lower psychological distress (b = −0.16, [-0.26, −0.06], τ*_b_* = 0.27, [0.17, 0.38]). Thus, had health satisfaction been compromised during the lockdown, people would have experienced greater psychological distress (see: Figure F1 and G1).

The lockdown did not reliably affect subjective health ratings (a = 0.04, [-0.02, 0.09], τ*_a_* = 0.07, [0.00, 0.21]), and so there was not a mediation effect of the lockdown on distress through subjective health ratings (me = −0.02, [-0.06, 0.03]). However, subjective health ratings predicted lower psychological distress (b = −0.48, [-0.68, −0.28], τ*_b_* = 0.69, [0.45, 0.92]). Thus, had subjective health ratings been compromised during the lockdown, people would have experienced greater psychological distress (see: Figure F2 and G2).

Similarly, at the population-level average, the lockdown did not reliably affect rumination (a = −0.03, [-0.08, 0.02], τ*_a_* = 0.12, [0.02, 0.27]), and so there was not a mediation effect of the lockdown on distress through rumination (me = −0.06, [-0.16, 0.03]). However, elevated rumination strongly predicted psychological distress (b = 1.53, [1.33, 1.72], τ*_b_* = 1.00, [0.81, 1.19]). Thus, had rumination increased during the lockdown, people would have experienced greater psychological distress (see: Figure F3 and G3).

At the population-level average, we find that the lockdown reduced fatigue (a = −0.21, [-0.26, −0.16], τ*_a_* = 0.11, [0.01, 0.29]), and that this reduction in fatigue indirectly reduced psychological distress (me = −0.19, [-0.27, −0.11]). Without such a reduction in fatigue, observed psychological distress (the total effect: *c′* = 0.44, [0.27, 0.60], τ*_c′_* = 0.22, [0.02, 0.57]) would have been substantially greater (c = 0.25, [0.08, 0.42]). Thus, rest during the lockdown was a powerful buffer against psychological distress. Figure F4 presents mediation results for fatigue (see: G4).

### Economic mediators of lockdown distress

We do not find a reliable effect of the lockdown on satisfaction with the economy (a = 0.06, [-0.07, 0.19], τ*_a_* = 0.24, [0.03, 0.54]), and so there was not a mediation effect of the lockdown on distress through satisfaction with the economy (me = 0.01, [-0.02, 0.05]). This is because satisfaction with the economy did not reliably predict psychological distress (b = −0.05, [-0.13, 0.02], τ*_b_* = 0.19, [0.08, 0.31]) (see: Figure F5 and G5).

There was not a reliable effect of the lockdown on satisfaction with standard of living (a = 0.01, [-0.08, 0.10], τ*_a_* = 0.17, [0.01, 0.42]), and so there was not a mediation effect of the lockdown on distress through satisfaction with standard of living (me = 0.01, [-0.03, 0.05]). However, satisfaction with standard of living predicted lower psychological distress (b = −0.13, [-0.24, −0.01], τ*_b_* = 0.30, [0.15, 0.45]). Thus, had satisfaction with standard of living been compromised during the lockdown, people would have experienced greater psychological distress (see: Figure F6 and G6).

There was not a reliable effect of the lockdown on satisfaction with future security (a = −0.10, [-0.22, 0.03], τ*_a_* = 0.32, [0.07, 0.61]), and so there was not a mediation effect of the lockdown on distress through satisfaction with future security (me = 0.01, [-0.06, 0.08]). However, satisfaction with future security reliably predicted lower psychological distress (b = −0.11, [-0.20, −0.02], τ*_b_* = 0.40, [0.28, 0.51]). Thus, had satisfaction with future security been compromised during the lockdown, people would have experienced greater psychological distress (see: Figure F7 and G7).

We find a reliable effect of the lockdown on business satisfaction (a = −0.32, [-0.44, −0.20], τ*_a_* = 0.69, [0.50, 0.93]); however, there was not a reliable mediation effect of the lock-down on distress through business satisfaction (me = −0.01, [-0.07, 0.06]). This is because business satisfaction did not reliably predict psychological distress (b = −0.04, [-0.14, 0.05], τ*_b_* = 0.16, [0.02, 0.31]). However, we caution that substantial declines in business satisfaction are evident in about a quarter of the population (see: Section A). Thus, although at the population-level average business satisfaction did not reliably predict psychological distress, we find signals of augmented distress among a sizable subgroup who fell in business satisfaction (see: Figure F8 and G8).

We do not find a reliable effect of the lockdown on social support (a = 0.03, [-0.03, 0.08], τ*_a_* = 0.10, [0.01, 0.26]), and so there was not a mediation effect of the lockdown on distress through social support (me = 0.00, [-0.05, 0.07]). However, social support reliably buffered people from psychological distress (b = −0.49, [-0.70, −0.27], τ*_b_* = 0.76, [0.48, 1.03]). Thus, had social support been compromised during the lock-down, people would have experienced greater psychological distress (see: Figure F9 and G9).

We do not find a reliable effect of the lockdown on social belonging (a = −0.03, [-0.08, 0.01], τ*_a_* = 0.07, [0.00, 0.18]), and so there was not a mediation effect for the lockdown on distress through social belonging (me = 0.02, [-0.02, 0.07]). However, social belonging reliably predicted lower psychological distress (b = −0.56, [-0.77, −0.36], τ*_b_* = 0.71, [0.50, 0.92]). Thus, had social belonging been compromised during the lockdown, people would have experienced more distress (see: Figure F10 and G10).

We find that the lockdown decreased satisfaction with personal relationships (a = −1.18, [-1.33, −1.03], τ*_a_* = 0.61, [0.34, 0.89]), and that satisfaction with personal relationships reliably predicted lower psychological distress (b = -0.16, [-0.24, −0.07], τ*_b_* = 0.27, [0.16, 0.38]); hence, there was a reliable mediation effect of the lockdown on distress through (dis)satisfaction with personal relationships (me = 0.20, [0.09, 0.32], (c = 0.25, [0.08, 0.42]). Indeed, the mediation effect here rendered the direct effect of lockdown on distress unreliable (c′ = 0.05, [-0.15, 0.24], τ′*_c_* = 1.30, [0.75, 2.15]). Thus, relationship dissatisfaction during the lock-down was a powerful instigator of psychological distress. Figure F11 presents mediation results for satisfaction with personal relationships (see: G11).

### Institutional trust mediators of lockdown distress

We find a reliable effect of the lockdown on satisfaction with the performance of government (a = 1.48, [1.36, 1.60], τ*_a_* = 0.27, [0.03, 0.62]), but not a mediation effect of the lock-down on distress through satisfaction with the performance of government (me = 0.10, [-0.03, 0.23]). This is because satisfaction with the performance of government did not reliably predict psychological distress (b = 0.07, [-0.01, 0.16], τ*_b_* = 0.16, [0.02, 0.34]) (see: Figure F12 and G12).

We find a reliable effect of the lockdown on trust in politicians (a = 0.39, [0.32, 0.47], τ*_a_* = 0.11, [0.01, 0.29]), but not a mediation effect of the lockdown on distress through trust in politicians (me = 0.01, [-0.05, 0.08]). This is because trust in politicians did not reliably predict psychological distress (b = 0.02, [-0.11, 0.16], τ*_b_* = 0.50, [0.30, 0.69]) (see: Figure F13 and G13).

We find a reliable effect of the lockdown on willingness to engage with the police (a = −0.16, [-0.21, −0.10], τ*_a_* = 0.24, [0.12, 0.41]), but not a mediation effect of the lockdown on distress through willingness to engage with the police (me = −0.03, [-0.10, 0.04]). This is because willingness to engage with the police did not reliably predict psychological distress (b = 0.00, [-0.21, 0.21], τ*_b_* = 0.63, [0.34, 0.89]) (see: Figure F14 and G14).

We find a reliable effect of the lockdown on trust in the police (a = 0.27, [0.21, 0.32], τ*_a_* = 0.13, [0.01, 0.32]), but not a mediation effect of the lockdown on distress through trust in the police (me = −0.05, [-0.11, 0.01]). This is because trust in the police did not reliably predict psychological distress (b = −0.18, [-0.37, 0.02], τ*_b_* = 0.23, [0.03, 0.47]) (see: Figure F15 and G15).

### Civic and community mediators of lockdown distress

We find a reliable effect of the lockdown on satisfaction with social conditions (a = 0.14, [0.02, 0.27], τ*_a_* = 0.26, [0.03, 0.56]), but not a mediation effect of the lockdown on distress through satisfaction with social conditions (me = 0.00, [-0.03, 0.04]). This is because satisfaction with social conditions did not reliably predict psychological distress (b = −0.03, [-0.12, 0.05], τ*_b_* = 0.13, [0.01, 0.28]) (see: Figure F16 and G16).

We find a reliable effect of the lockdown on national identity (a = 0.09, [0.05, 0.13], τ*_a_* = 0.21, [0.10, 0.38]), but not a mediation effect of the lockdown on distress through national identity (me = 0.01, [-0.04, 0.07]). This is because national identity did not reliably predict psychological distress (b = 0.21, [-0.08, 0.54], τ*_b_* = 0.54, [0.13, 0.92]) (see: Figure F17 and G17).

We find a reliable effect of the lockdown on patriotism (a = 0.24, [0.20, 0.28], τ*_a_* = 0.18, [0.07, 0.33]), but not a mediation effect of the lockdown on distress through patriotism (me = 0.08, [-0.01, 0.19]). However, interestingly, patriotism predicted greater psychological distress (b = 0.27, [0.00, 0.54], τ*_b_* = 0.99, [0.37, 1.36]), though with substantial individual-level variability (τ*_b_* = 0.99, [0.37, 1.36], see Section A) (see: Figure F18 and G18).

We find a reliable effect of the lockdown on sense of neighbourhood community (a = 0.25, [0.18, 0.33], τ*_a_* = 0.18, [0.02, 0.43]), and a reliable mediation effect of the lockdown on distress through a sense of neighbourhood community (me = −0.05, [-0.11, 0.00]). Because sense of neighbourhood community reliably predicts lower psychological distress (b = −0.13, [-0.26, −0.01], τ*_b_* = 0.28, [0.12, 0.45]), the increase in a sense of neighbourhood community during the lockdown helped to buffer people from greater psychological distress. Figure F19 presents mediation results for sense of neighbourhood community (see: G19).

## Discussion

### (1) Rest during the lockdown diminished psychological distress

Fatigue reduction powerfully mitigated psychological distress during New Zealand’s stringent lockdown in March/April 2020. To clarify the mechanism of this effect, we graph the relationship between the fitted values of the model for the ‘a’ path (X→M) in Figure 1.i. Here, we find an expected reduction in fatigue across the population during the lockdown. We next graph the relationship between the fitted values of the model for the ‘b’ path (M→Y) in Figure 1.ii. Here, we find expected increases in psychological distress for increasing levels of fatigue. Although not all New Zealanders were afforded relief from fatigue during lockdown, such as essential workers and parents with children (Twenge & Joiner, 2020), were it not for a general reduction in fatigue across most of the population, we estimate that psychological distress during the lockdown would have been 1.74 times greater (Fatigue c = 0.25 [0.08, 0.42], Fatigue cp = 0.44 [0.27, 0.60]).

**Figure 1.**
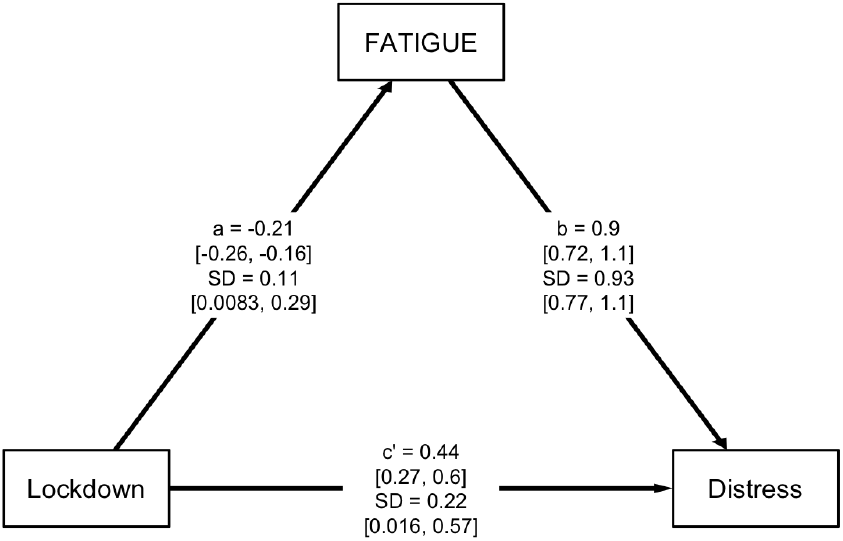
Panel (i) presents the fitted values for the ‘a’ path X→M, indicating that the lockdown caused people to feel less fatigue. Panel (ii) presents the fitted values for the ‘b’ path (M→Y), indicating that psychological distress is strongly sensitive to levels of fatigue. The model indicates that psychological distress would have been 1.74 times greater had lockdown not reduced fatigue.

The association between fatigue and psychological distress has long been observed (Pawlikowska et al., 1994; Thorndike, 1900); however, longitudinal dynamics of fatigue and mental health across a population remain poorly understood. The powerful psychological distress buffering that we observe from reduced fatigue suggests the systematic study of rest is likely important for pandemic mental health research, and for public mental health more generally. (On measures, see: Dittner et al., 2004).

### (2) Stable health satisfaction during the lockdown spared psychological distress

Despite the pandemic health uncertainties confronting New Zealand during the March/April 2020 lockdown, health satisfaction and subjective health ratings did not decline, and levels of rumination were not elevated. Statistical models reveal that had health satisfaction and subjective health declined, and rumination elevated, psychological distress would have risen (see: Figure B1). These results indicate that New Zealand’s initial sheltering-in-place lockdown sustained mental health because it afforded physical health security. Generalising, our findings add credibility to Qiu et al. (2020)’s and Twenge and Joiner (2020)’s speculation that health concerns drove psychological distress during the early COVID-19 pandemic in settings where personal infectious disease risks were high.

### (3) Stable standards of living during the lockdown spared psychological distress

Levels of business satisfaction dropped from pre-pandemic levels during New Zealand’s March/April lockdown. That business satisfaction declined is, on reflection, not surprising, considering all non-essential businesses were closed to the public for an indefinite period. Among a sizeable subgroup of the population this drop in business satisfaction provoked psychological distress.

Importantly, however, satisfaction with standard of living and future security were maintained at pre-pandemic levels. This stability is both surprising and important. It reveals that even in a setting of pervasive pandemic economic uncertainties and forced business closures, maintaining satisfaction with standard of living and a sense of future security is possible. Moreover, the strong sensitivity of psychological distress to economic security implies that had economic concerns become elevated, psychological distress during the lockdown would also have become more elevated (see: Figure B2). We infer that New Zealand’s rapid and robust administration of economic relief to citizens benefited mental health during the lockdown because it afforded economic security.

### (4) The lockdown strained personal relationships and this strain accounts for most of the psychological distress that people experienced

Despite requirements for sheltering-in-place, social support and social belonging were conserved during New Zealand’s stringent March/April pandemic lockdown. We quantitatively demonstrate that were these parameters not conserved at pre-pandemic levels during the lockdown, psychological distress would have been substantially worse (see: Figure B3). This result supports Van Bavel et al. (2020)’s theory that maintaining social connection when physically distancing protects mental health.

However, New Zealand’s shelter-in-place lockdown exacted a toll on satisfaction with personal relationships. Indeed, the psychological distress that New Zealanders experienced during lockdown mostly arose from dissatisfaction with personal relationships (see: Figure 3).

**Figure 2.**
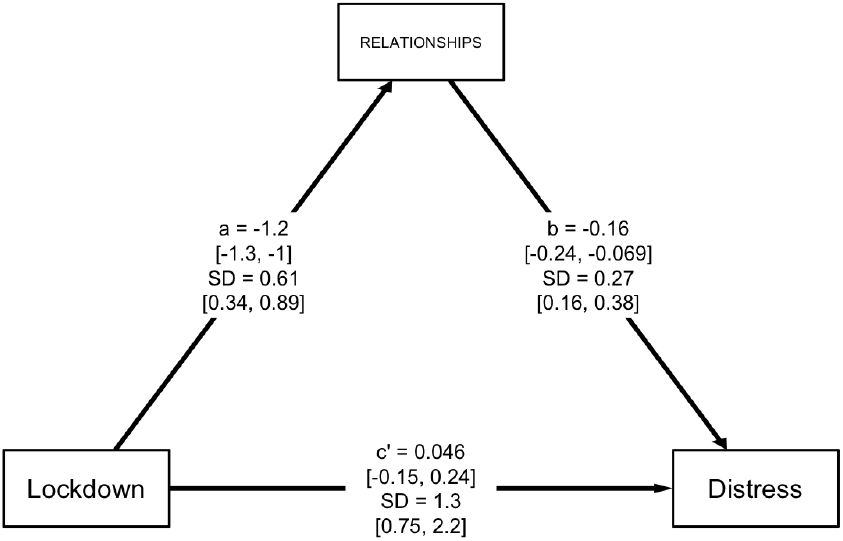
Panel (i) presents the fitted values for the ‘a’ path (X→M), indicating that the lockdown caused people to feel less satisfaction with their personal relationships. Panel (ii) presents the fitted values for the ‘b’ path (M→Y), indicating that lower satisfaction with personal relationships increases psychological distress. Indeed, the mediation effect of relationship dissatisfaction (me = 0.20, [0.09, 0.32]) was strong enough to render the direct effect of the lockdown on psychological distress unreliable (c′ = 0.05, [-0.15, 0.24]). This finding implies that relationship dissatisfaction was a powerful engine of psychological distress during New Zealand’s early COVID-19 lockdown.

**Figure 3.**
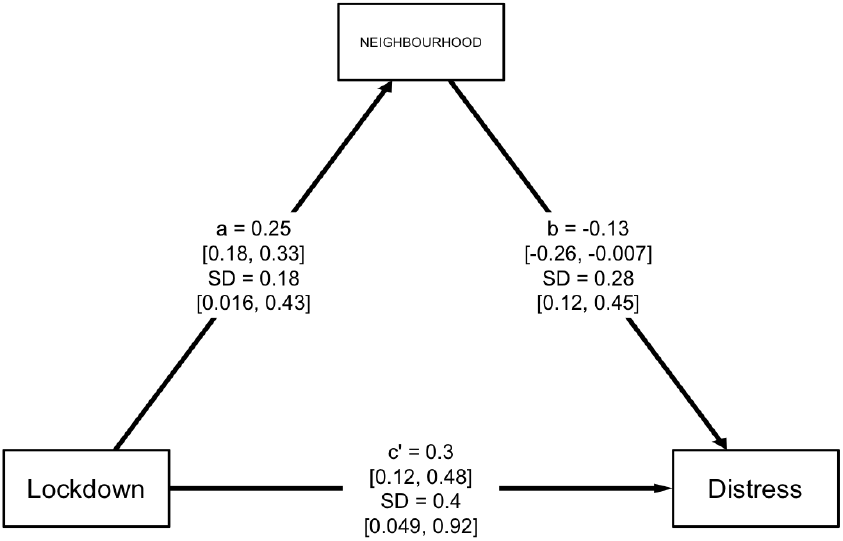
Panel (i) presents the fitted values for the ‘a’ path (X→M), indicating that the lockdown caused people to feel greater satisfaction with the New Zealand government. However, panel (ii) presents the fitted values for the ‘b’ path (M→Y), indicating that in New Zealand psychological distress is not generally sensitive to levels of satisfaction with the government’s performance. In a setting such as New Zealand’s, it is not necessary to shift a population’s satisfaction with the government to prevent elevation in pandemic distress.

The lockdown could have challenged personal relationships in a number of ways. One possibility is that the lockdown hindered close connections with friends and family outside of the home (Van Bavel et al., 2020). Yet, if this were the primary reason for the declines in satisfaction with personal relationships then the same negative effects would have emerged with social support and belonging.

Pietromonaco and Overall (2020) theorise that confinement in the home not only reduced people’s ability to control personal space, but required them to share the increased difficulties in balancing work, childcare and household labour. By narrowing personal networks, lockdown also amplified the risk of stress contagion and the burden of support for those in the home. All of these pressures are established risks to psychological health (Pietromonaco & Overall, 2020).

Our finding supports Pietromonaco and Overall (2020)’s theory that a stringent pandemic lockdown strains personal relationships, and furthermore reveals that such strains led to greater psychological distress. However, the present dataset does not allow us to infer which features of romantic relationship strain caused psychological distress to increase during lockdown, a matter for future investigations.

### (5) Greater satisfaction with the performance of the government did not reduce psychological distress

Despite a strong increase in satisfaction with the government, we do not observe a mediation effect of satisfaction with the government on psychological distress. This is because satisfaction with the government is not causally related to psychological distress. In New Zealand’s democracy, where 50– 60% of the population differ in their attitudes to major political parties, it is unsurprising that satisfaction with the government’s performance does not predict psychological distress. We cannot infer whether this finding generalises to countries where levels of satisfaction with the performance of the government are much lower.

### (6) During the lockdown an elevated sense of neighbourhood community reduced psychological distress

Consistent with Sibley, Greaves, et al. (2020)’s theory, we observe that an elevated sense of neighbourhood community contributed to reducing pandemic distress (see: Figure 4). Notably, the relationship between civic sensibilities and distress is weaker for abstract national orientations than it is for neighbourhood orientations.

**Figure 4.**
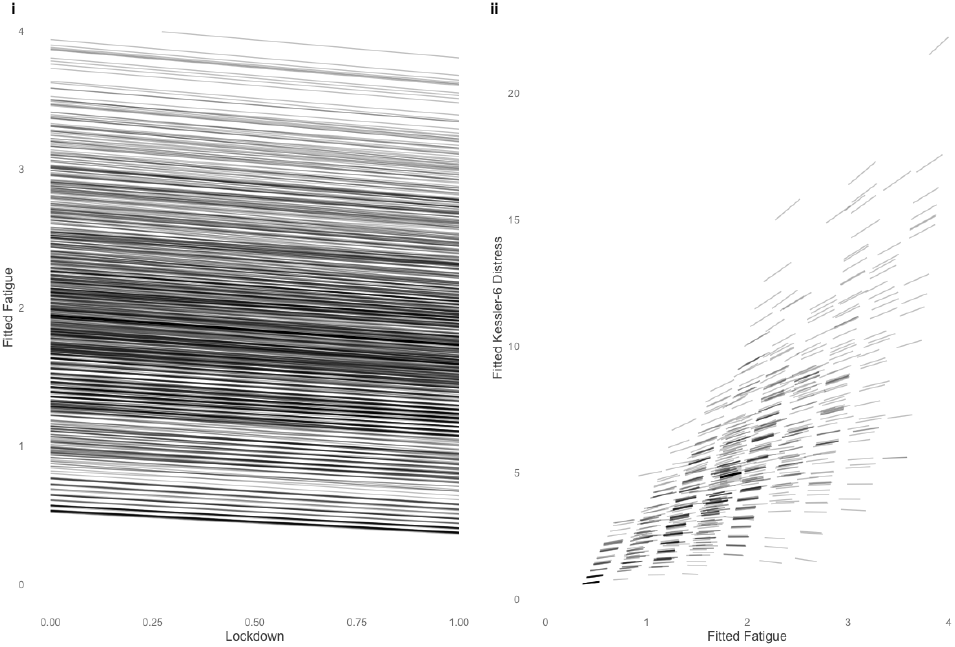
Panel (i) presents the fitted values for the ‘a’ path (X→M), indicating that the lockdown caused people to feel a greater sense of neighbourhood community. Panel (ii) presents the fitted values for the ‘b’ path (M→Y), indicating elevated neighbourhood community predicts lower psychological distress. This result suggests that in a setting such as New Zealand’s, improving a sense of neighbourhood community reduces pandemic psychological distress.

### (7) Key inference: at the onset of a lethal pandemic, the public health benefits of infectious disease containment extend to mental health

The late onset of the COVID-19 pandemic in New Zealand gave the country’s public health response a head start (see Appendix C). A cross-sectional study of Italy’s lockdown of April 10*^th^*–13*^th^* 2020 reveals substantial population-level distress (Moccia et al., 2020); however, the pandemic struck Italy early and with tragically high mortality. From the vantage point of early disease containment, it is relative timing that counts. Our national longitudinal study of pandemic distress dynamics leverages pre-pandemic responses to quantitatively demonstrate that there is a parallel benefit of infectious disease containment for psychological health. Why was distress only moderately elevated during New Zealand’s early pandemic in March/April? The answer is that New Zealand was in a strict and economically supportive pandemic lockdown. We find that New Zealand’s public health response tended to maintain New Zealanders’ sense of health and financial security, as well as their sense of social support and belonging. Additionally, we find that had these parameters been compromised, psychological distress would have been much worse (see: Figure B1). Moreover, we observe that an elevated sense of community played a small role in preserving residents mental health, and that a reduction in fatigue – rest – played a considerable role (see: Figure 1). The causal effects of fatigue and rest on psychological distress is an important horizon at the boundary of psychological science and public health. Importantly, sheltering-in-place strained personal relationships, and it was this strain that accounted for much of the distress that New Zealanders experienced during the country’s sheltering-in-place pandemic lockdown in March/April 2020.

Overall, New Zealand’s avoidance of the greater psychological distress found in other countries during the early COVID-19 pandemic reveals that the emergence of a new and lethal infectious disease need not fate a society to severe mental health burdens. Whether these benefits extend to subsequent pandemic lockdowns, and whether lockdowns generally sustain mental health over the long-term is presently unclear. Long-term mental health trajectories rely on an interplay of long-term social, health, and economic trajectories. These are questions of contemporary speculation and debate which we cannot resolve here (see: McKibbin and Fernando, 2020; Wang and Tang, 2020). On the other hand, it has long been understood that strict infectious disease containment benefits public health (Soper, 1919). Our findings are important to current and future pandemic planning because they quantitatively clarify fundamental psychological mechanisms through which stringent infectious disease containment strategies sustain mental health when lethal pandemics first strike.

## Data Availability

A de-identified dataset containing the variables analysed in this manuscript is available upon request from the corresponding author, or any member of the NZAVS advisory board for the purposes of replication or checking of any published study using NZAVS data. The code for all analyses and graphs is available on the Open Science Framework at: https://osf.io/5gza8/. Contacts for the NZAVS board: https://www.psych.auckland.ac.nz/en/about/new-zealand-attitudes-and-values-study/NZAVS-research-group.html

## Future Research

The limitations of our study suggest following opportunities for future research.

1. Pandemic distress mechanisms are potentially dynamic, cumulative, and nonlinear. Here, however, we restrict inferences about mechanisms of mental health to the onset of a lethal pandemic. We use data originally collected for Sibley, Greaves, et al. (2020)’s matched propensity-score investigation of attitudes and well-being during the first 18 days of New Zealand’s pandemic. These data cannot clarify long-term pandemic distress dynamics. As longer windows of data become available, future research should investigate these dynamics.
2. The present study takes the form of a natural experiment, with the control condition consisting of within-participant baselines obtained from the previous year. However, clearly, the random assignment of New Zealanders to pandemic conditions resembling Wuhan, Lombardy, New York City, or elsewhere is neither possible nor desirable. Our statistical models are able to quantitatively predict how much worse distress would have been had parameters important to mental health been compromised (see: Figures B1–B5). Additionally, these models allow disentangling the effects of the pandemic lockdown from the effects of health and economic uncertainties owing to the pandemic more generally. For example, because relationship dissatisfaction occurred in the absence of health and business concerns, we were able to infer that distress from relationship dissatisfaction arose from the demands of sheltering-in-place. However, we cannot estimate how much the mediation parameters themselves would have changed had the early pandemic taken a different form. Such an inference would rely on comparative panel data which is not presently available.
3. Relatedly, fundamental questions about the mechanisms of psychological distress in other countries require additional assumptions that our data cannot directly address. We cannot, for example, infer that people in other countries would have responded similarly to New Zealanders had they experienced New Zealand’s “go-hard, go-early” pandemic response.

To address these limitations, we recommend that national longitudinal health studies include indicators of psychological distress (we recommend the Kessler-6), as well as locally validated indicators for subjective health concerns, economic concerns, personal relationships, institutional trust, and civic sensibilities.

### Declarations

The New Zealand Attitudes and Values Study is supported by a grant from the Templeton Religion Trust (TRT0196). The funders had no role in any aspect of this study or the decision to publish. The authors declare no competing interests.

### Notes

JB did the analysis and wrote the first draft with SP. CS leads the New Zealand Attitudes and Values Study and managed data collection. All authors contributed substantially to the research, writing, or data collection for this study.

## Appendix A Investigation of Individual Differences in Psychological Distress

### Individual differences in health mediators

The coefficients for the four health dimensions are graphed in Figure A1a.i–A1d.i. In addition to the population-level coefficients, we include individual-level estimates for a, b, and me parameters in Figure A1 (rows ii-iv); these individual-level estimates afford a window into patterns of individual difference. There is more uncertainty in estimates of the lockdown’s direct effect on health satisfaction (τ*_a_* = 0.15, [0.01, 0.42], see: Figure A1a.ii and Figure F1), than its effect on subjective health ratings (i.e., short form health) (τ*_a_* = 0.07, [0.00, 0.21], see: Figure A1b.ii and Figure F2).

There were comparable magnitudes of individual-level variation in the lockdown’s effect on rumination (τ*_a_* = 0.12, [0.02, 0.27], see: Figure A1c.ii and Figure F3). Furthermore, we observe structured variation at the individual-level mediation estimates for rumination, where 6% were expected to be lower than −0.25 points in distress response (τ*_me_* = −0.06 [-0.44, 0.28]). The relief from diminished rumination at the lower end of distress is evident in Figure A1c.iv.

This pattern we observe for the effect of diminished rumination on distress reduction is consistent with the pattern we observe for the effect of diminished fatigue on distress reduction; however, as noted, mediation effects for fatigue were reliable (me = −0.19, [-0.27, −0.11]). Similar to rumination, the mediation effect for fatigue exhibited a downward pull at the lower end of the distress response spectrum (τ*_me_* = −0.19 [-0.55, 0.12], see: Figure A1d.ii and Figure F4).

### Individual differences in economic mediators

In Figure A2 we present the results for the four economic dimensions; to clarify individual differences we also include individual-level estimates for the mediation parameters.

We observe individual-level variation in the effects of lockdown on future security τ*_a_* = 0.32, [0.07, 0.61]). Here, 15.2% of the individual-level variances in future security were estimated to have fallen by more than 0.25 points, and 0.1% lower than 0.5 points, a rare but steep decline. We estimated substantial increases in distress greater than 0.25 points to arise from loss of future security in 0.3% of the population (see: Figure A2c.iv.). Though such effects were rare, they are not trivial.

We observe substantial individual-level variation in the effects of lockdown on business satisfaction τ*_a_* = 0.69, [0.50, 0.93]), presented in Figure A2d.ii. The minor elevation of business satisfaction in a minority of the population masked greater declines in business satisfaction, that were particularly severe at the lower end. Whereas only 0.002% of the individual-level variance in τ*_a_* for business satisfaction increased by greater than 0.25 points, 57.2% of the individual-level variances were dropped by more than 0.25 points, and 32.1% dropped by more than 0.5 points in business satisfaction (see: Figure A2d.ii). While there is uncertainty at the margins of the business satisfaction mediation effect, only 0.002% of the individual-level variances exhibited declines of more than 0.2 points, and 1% exhibited increases in excess of 0.2 points (see: Figure A2d.iv). However, business satisfaction does not strongly or reliably affect psychological distress, which suggests business satisfaction would constrain the mediation effect even if lockdown were to have had a stronger effect on business satisfaction (b = −0.04, [-0.14, 0.05]).

### Individual differences in social relationship mediators

In Figure A3 we present the results for the three social relationship parameters; to clarify individual differences we also include individual-level estimates for the mediation parameters (rows ii–iv).

We observe strong sensitivity of distress to both social support τ*_b_* = 0.76, [0.48, 1.03]), and social belonging (τ*_b_* = 0.71, [0.50, 0.92], see: Figure A3a.iii and A3b.iii), yet little movement in these parameters during lockdown (social support, τ*_a_* = 0.10, [0.01, 0.26]; social belonging, τ*_a_* = 0.07, [0.00, 0.18]). Thus, consistent with speculation in Van Bavel et al. (2020), these results suggest that had social support or social belonging suffered, New Zealanders would have experienced stronger challenges to mental health across the population.

At the individual-level, we observe substantial variation in the effect of lockdown on relationship (dis)satisfaction (τ*_a_* = 0.61 [0.34, 0.89]). As evident in Figure A3d.ii, such variation was greater at the low end of the distress response spectrum, with 16.9% of individual-level estimates falling by more than 1.5 points, and 2% falling by more than 2 points. As evident in Figure A3d.iv, there were strong mediation effects at the high end of the distress response spectrum, where 8.2% of the individual-level estimates exceeded a 0.5 point expected increase in psychological distress, and 1% exceeded a 1 point expected increase. Given the relative stability of health and economic concerns that we previously observed, we infer that the increase in relationship dissatisfaction arose from the requirement to shelter-in-place with domestic cohabitants. Though people felt safe in their health, future security, and standard of living, their personal relationships suffered.

**Figure A1.**
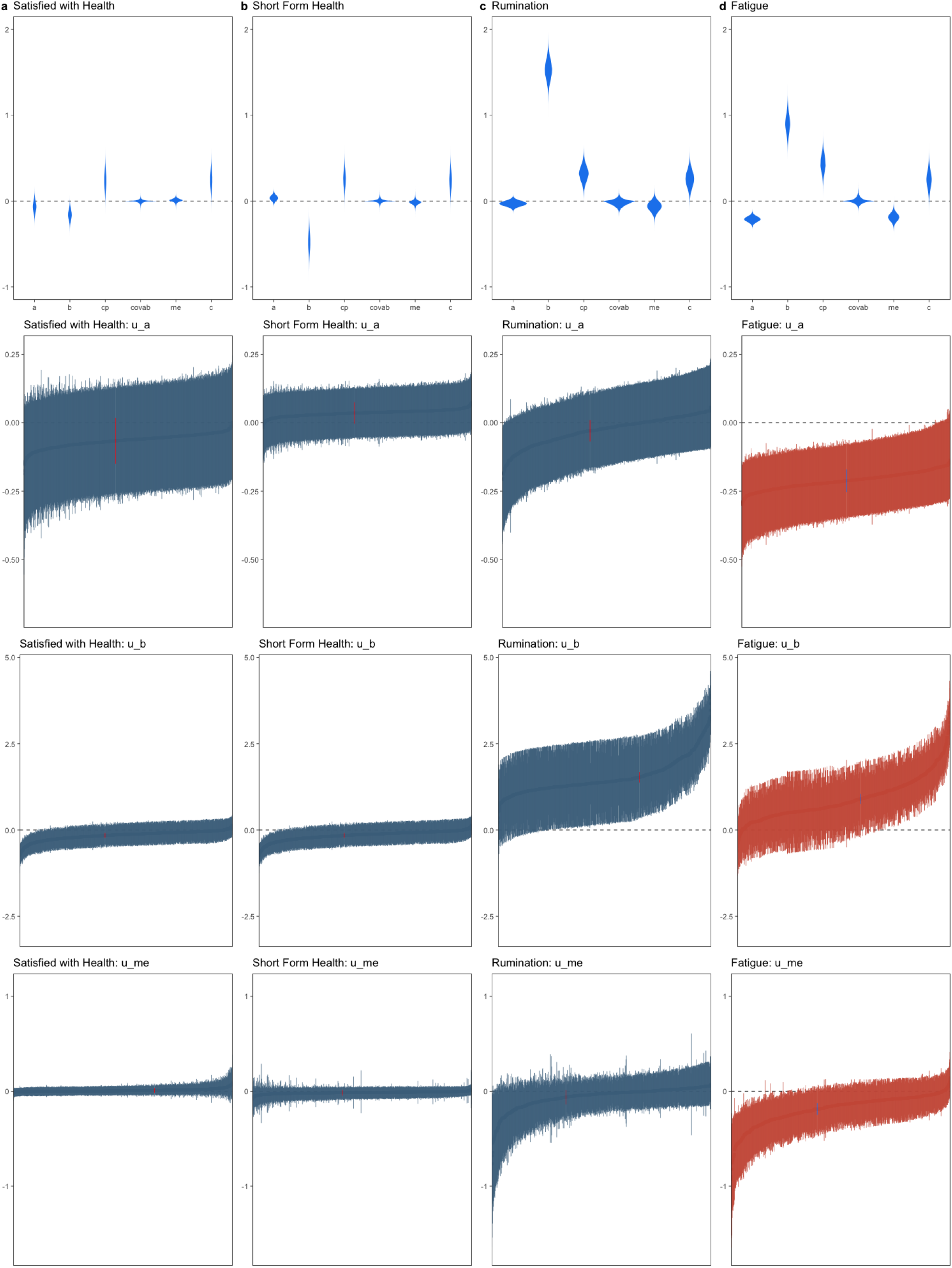
Individual differences in mediation mechanisms for heath parameters reveals substantial relief from psychological distress from diminished rumination at the margins of response, consistent with evidence for strongly diminished fatigue at the margins of response.

**Figure A2.**
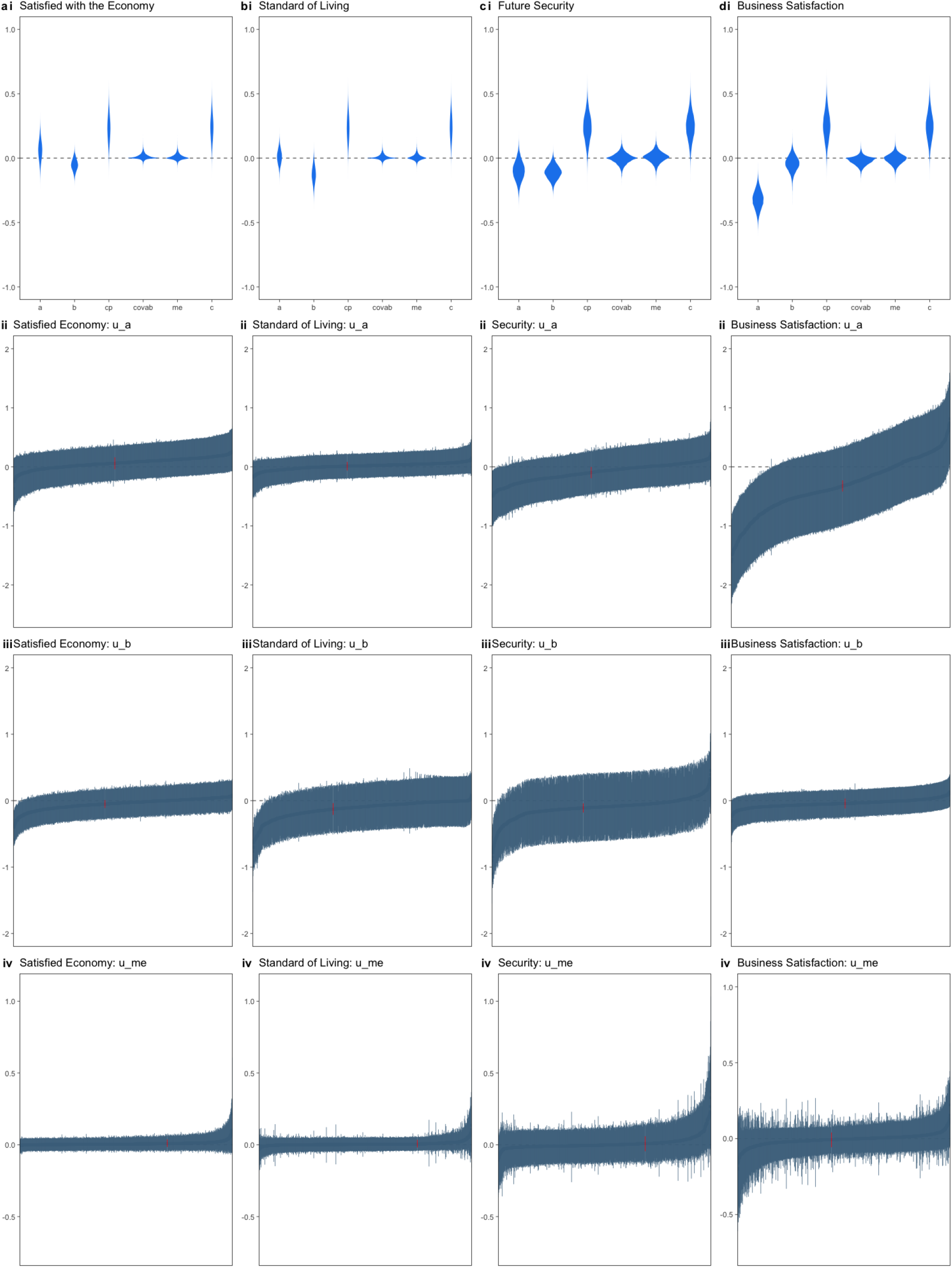
Individual differences in mediation mechanisms for economic parameters reveals variability in the effects of the lockdown on future security and business satisfaction, with some evidence for increased distress mediated by loss of future security at margins of the population average.

**Figure A3.**
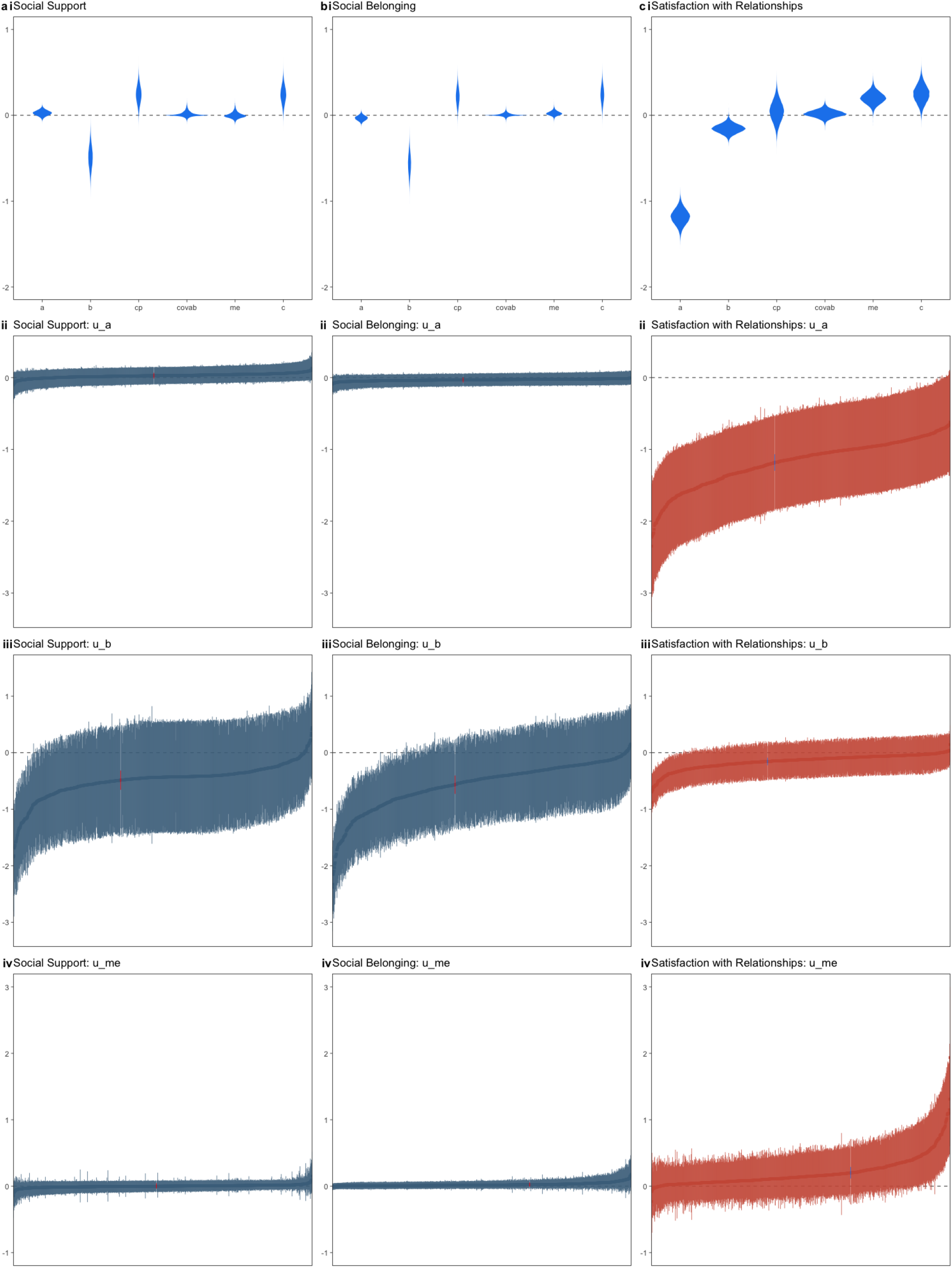
Individual differences in mediation mechanisms for relationship parameters reveals mediation effects were driven by extreme responses in the upper quartile of the ‘me’ estimates.

### Individual differences in institutional trust mediators

In Figure A4 we present the results for the four institutional trust dimensions; to clarify individual differences we also include individual-level estimates for the mediation parameters (rows ii–iv).

We observe substantial individual variation in the relationship between trust of politicians on psychological distress τ*_b_* = 0.50, [0.30, 0.69], and willingness to engage with the police on psychological distress (τ*_b_* = 0.63, [0.34, 0.89], see: Figure A4b.iii and c.iii). New Zealanders, then, differ in how these two dimensions of institutional distrust affect psychological distress; understanding these differences is a matter for future investigations.

### Individual differences in civic community mediators

In Figure A5 we present the results for the four civic community dimensions; to clarify individual differences parameters, we also include individual-level estimates (rows ii–iv).

We observe substantial and structured individual variability in the relationship between lockdown and both national identity and patriotism (national identity τ*a* = 0.21, [0.10, 0.38]; patriotism τ*a* = 0.18, [0.07, 0.33], see: Figures A5b.ii and A5c.ii). We also find substantial variability in the relationship of these two parameters to psychological distress (national identity, τ*_b_* = 0.54, [0.13, 0.92]; patriotism τ*_b_* = 0.99 [0.37, 1.36], see: Figures A5b.iii and A5c.iii), corresponding to variability in the individual-level mediation effects for these parameters, see Figure A5b.iv and A5c.iv. In the case of patriotism, whereas 1% of individual-level estimates were lower than −0.25 points in psychological distress, 6% were greater than 0.25 points in psychological distress. Why the increase in patriotism from lockdown should increase psychological distress for that segment of the New Zealand population is a matter for future investigation. By contrast, the expected benefits of increased neighbourhood community were consistent with predictions.

**Figure A4.**
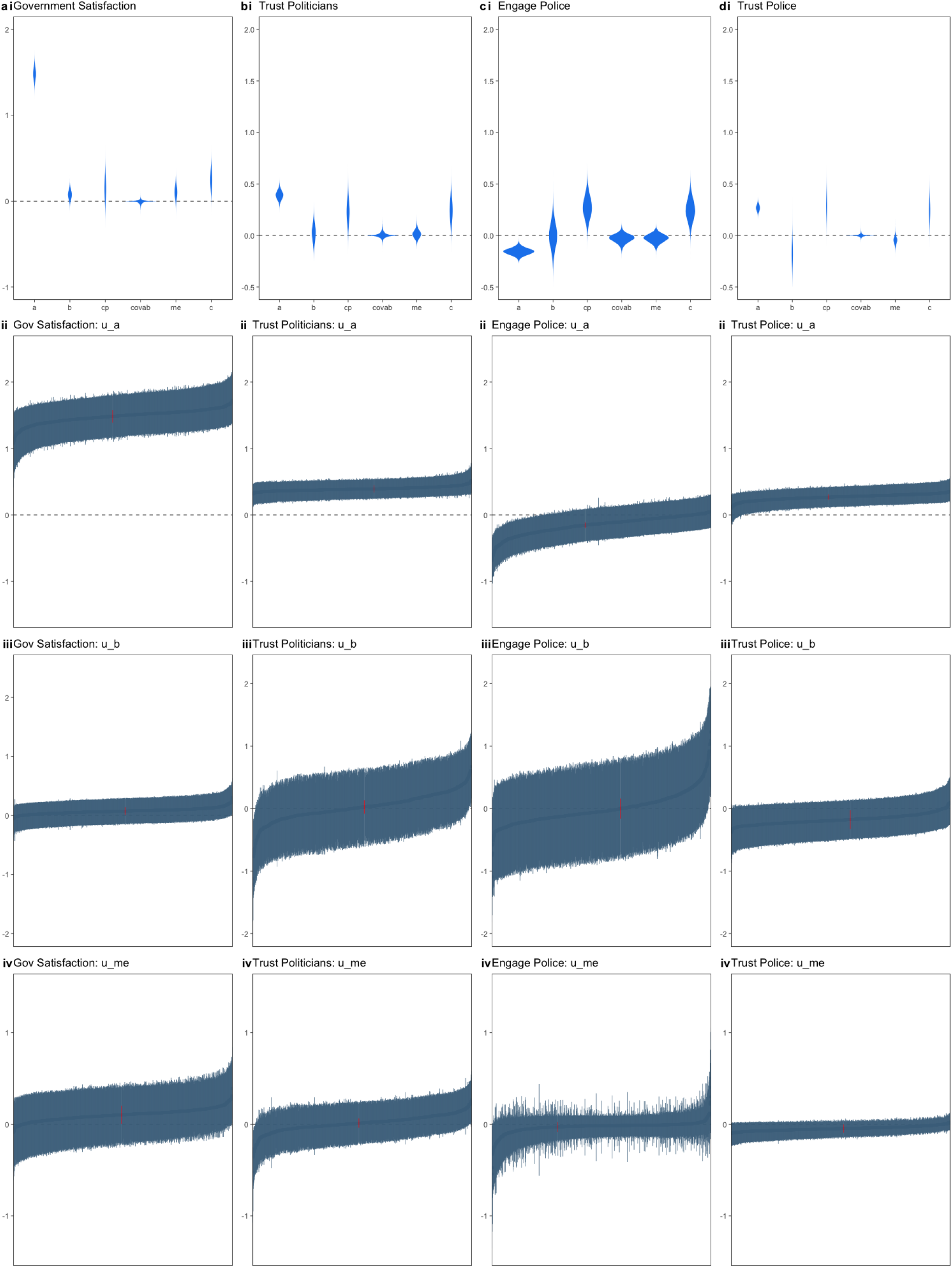
Individual differences in mediation mechanisms for institutional trust parameters reveals structured variation in trust of politicians and willingness to engage police.

**Figure A5.**
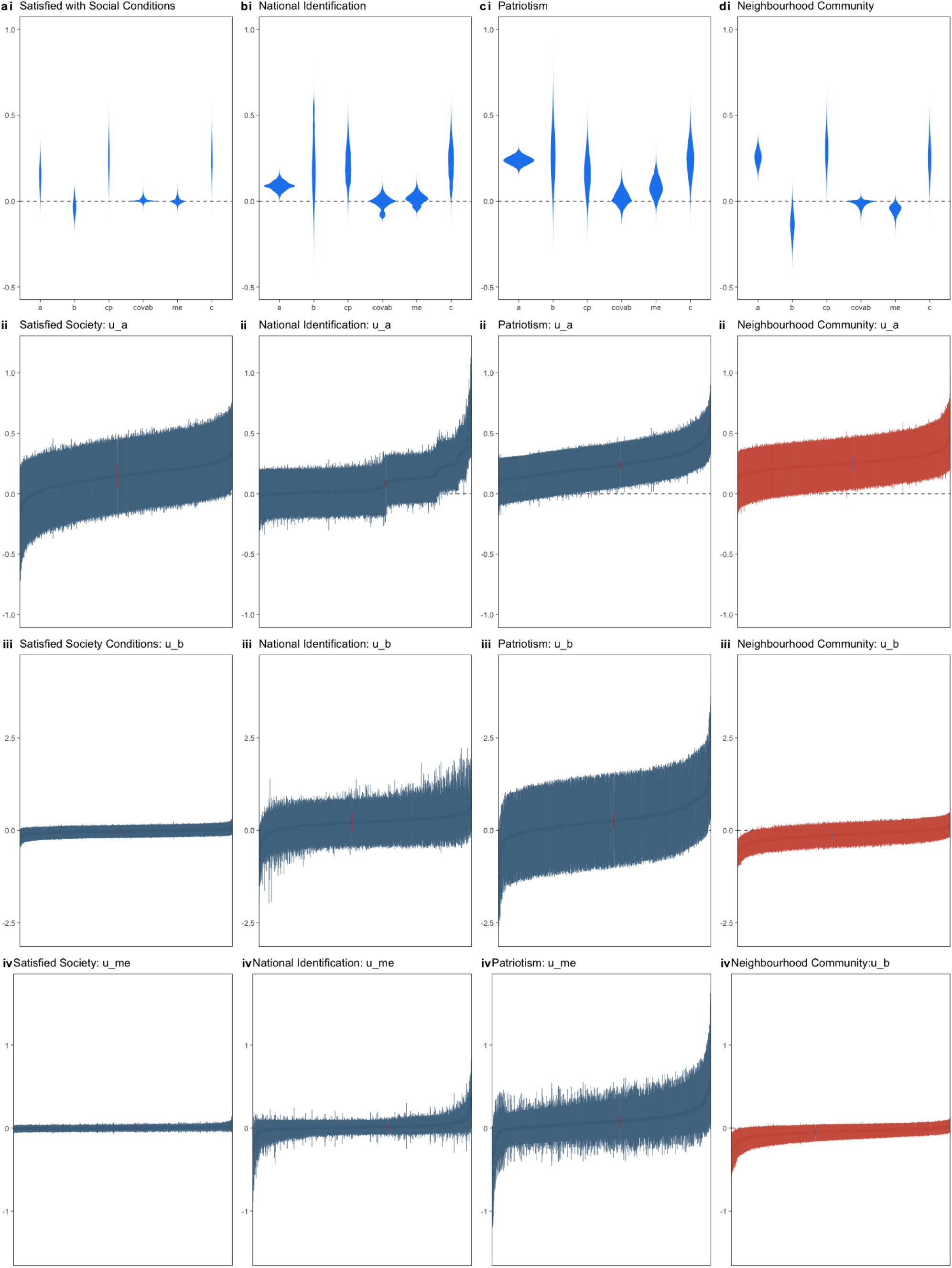
Individual differences in mediation mechanisms for civic community parameters reveals variation in the mediation effects of lockdown for these parameters.

## Appendix B Predictive Graphs

**Figure B1.**
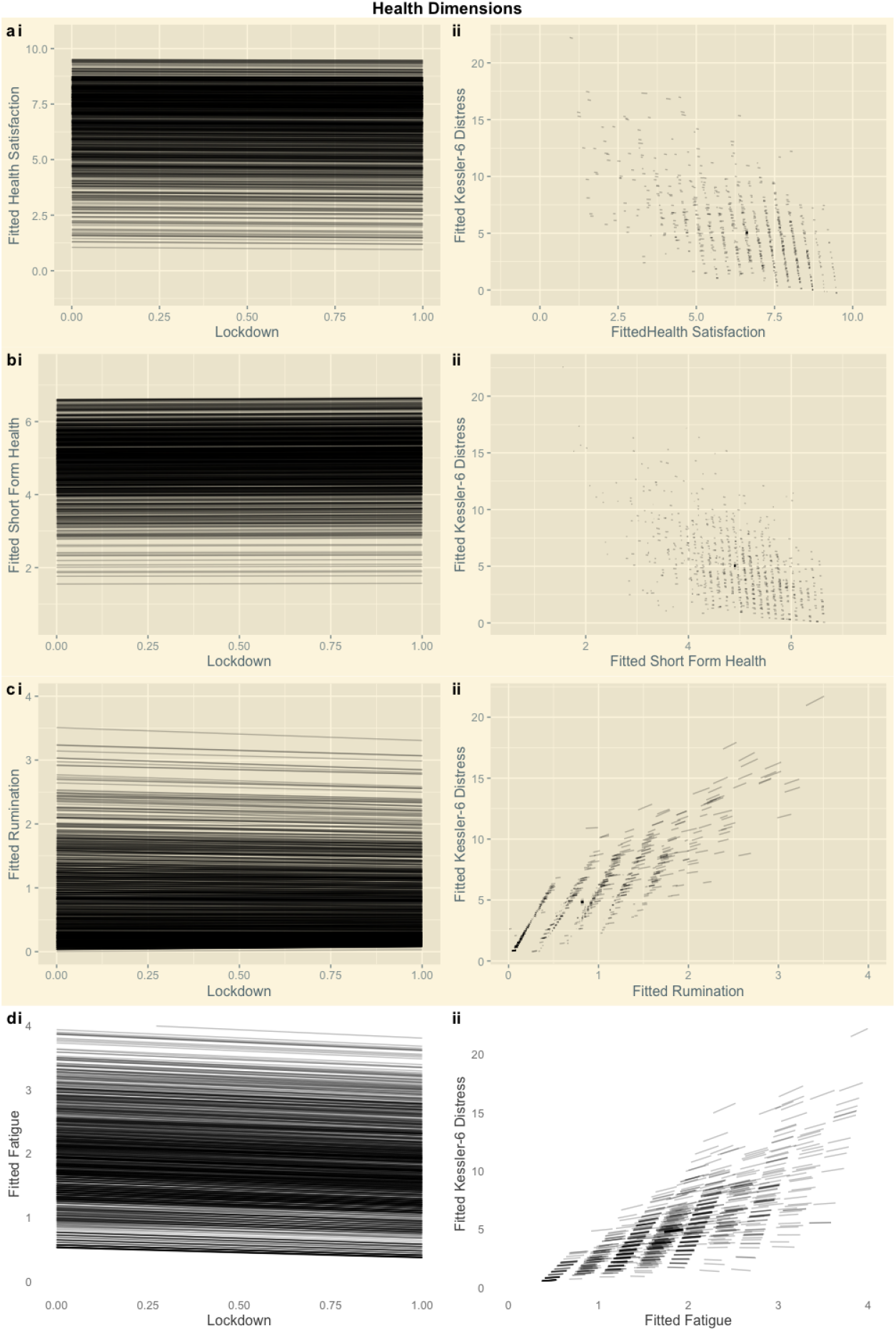
Predictive Plots: Personal Health. Panel (i) presents the fitted values for the ‘a’ path (X→M). Panel (ii) presents the fitted values for the ‘b’ path (M→Y). We do not observe reliable mediation effects for the health indicators. However, we observe strong relationships between health indicators and psychological distress (the ‘b’ paths), which reveal that if health had been compromised during the lockdown then New Zealanders would have experienced greater psychological distress. The white panels describe the ‘a’ and ‘b’ paths for the reliable mediation effect of lockdown on psychological distress through fatigue reduction. Specifically, the lockdown decreased fatigue, and furthermore, because fatigue elevation predicts psychological distress, there was a buffering mediation effect.

**Figure B2.**
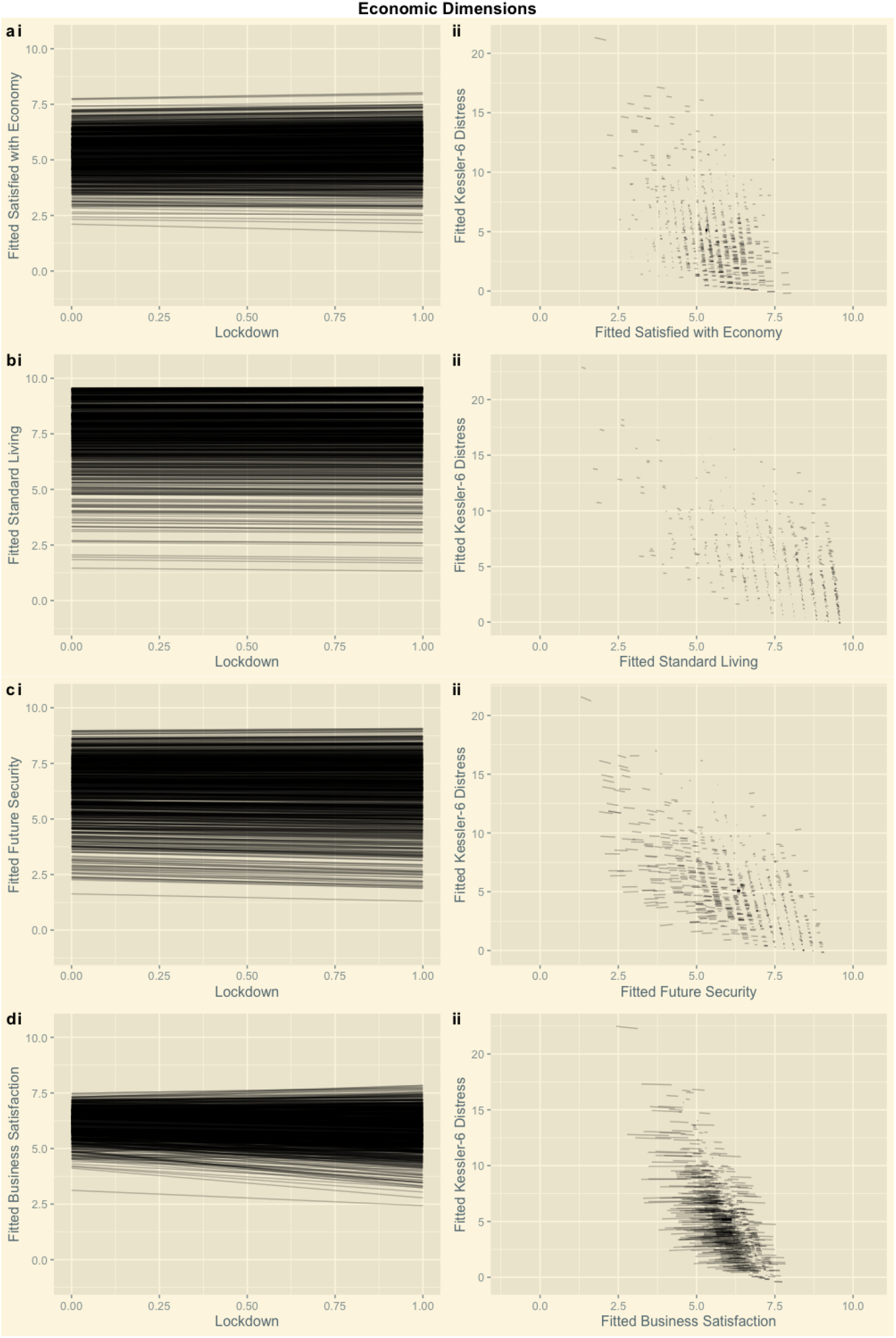
Predictive Plots: Economic Concerns. Panel (i) presents the fitted values for the ‘a’ path X→M. Panel (ii) presents the fitted values for the ‘b’ path (M→Y). We do not observe reliable mediation effects for economic concerns. However, we observe strong relationships between economic concerns and psychological distress (the ‘b’ paths), which reveal that if economic concerns had been elevated during the lockdown then New Zealanders would have experienced greater psychological distress.

**Figure B3.**
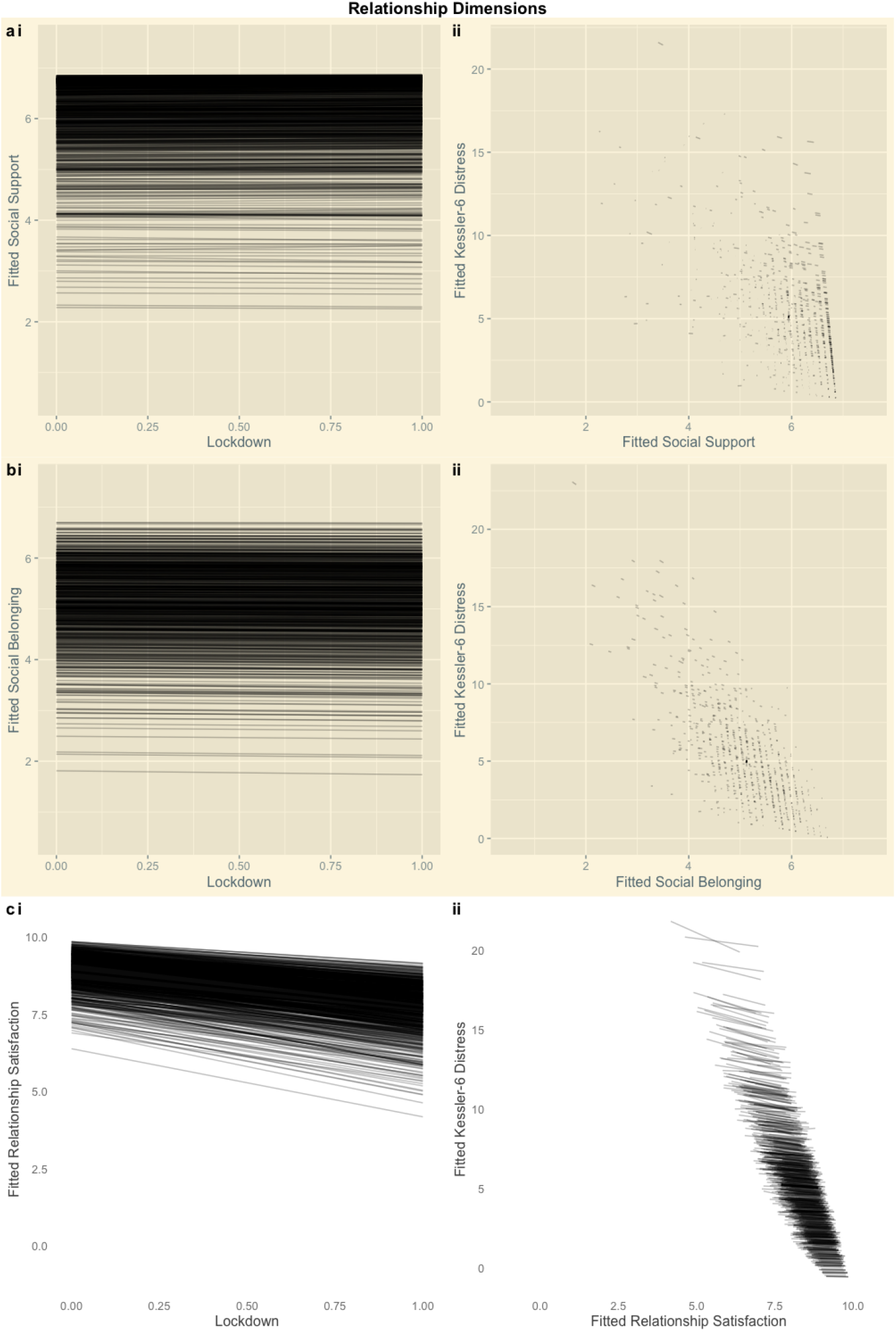
Predictive Plots: Social Relationships. Panel (i) presents the fitted values for the ‘a’ path (X→M). Panel (ii) presents the fitted values for the ‘b’ path (M→Y). We do not observe reliable mediation effects for social belonging and social support. However, we observe strong relationships between these parameters and psychological distress (the ‘b’ paths), which reveal that if social belonging or social support had been compromised during the lockdown then New Zealanders would have experienced greater psychological distress. The white panels describe the ‘a’ and ‘b’ paths for the reliable mediation effect of the lockdown on psychological distress through relationship dissatisfaction. Specifically, the lockdown decreased relationship satisfaction, and furthermore, because relationship dissatisfaction elevated psychological distress, there was a mediation effect.

**Figure B4.**
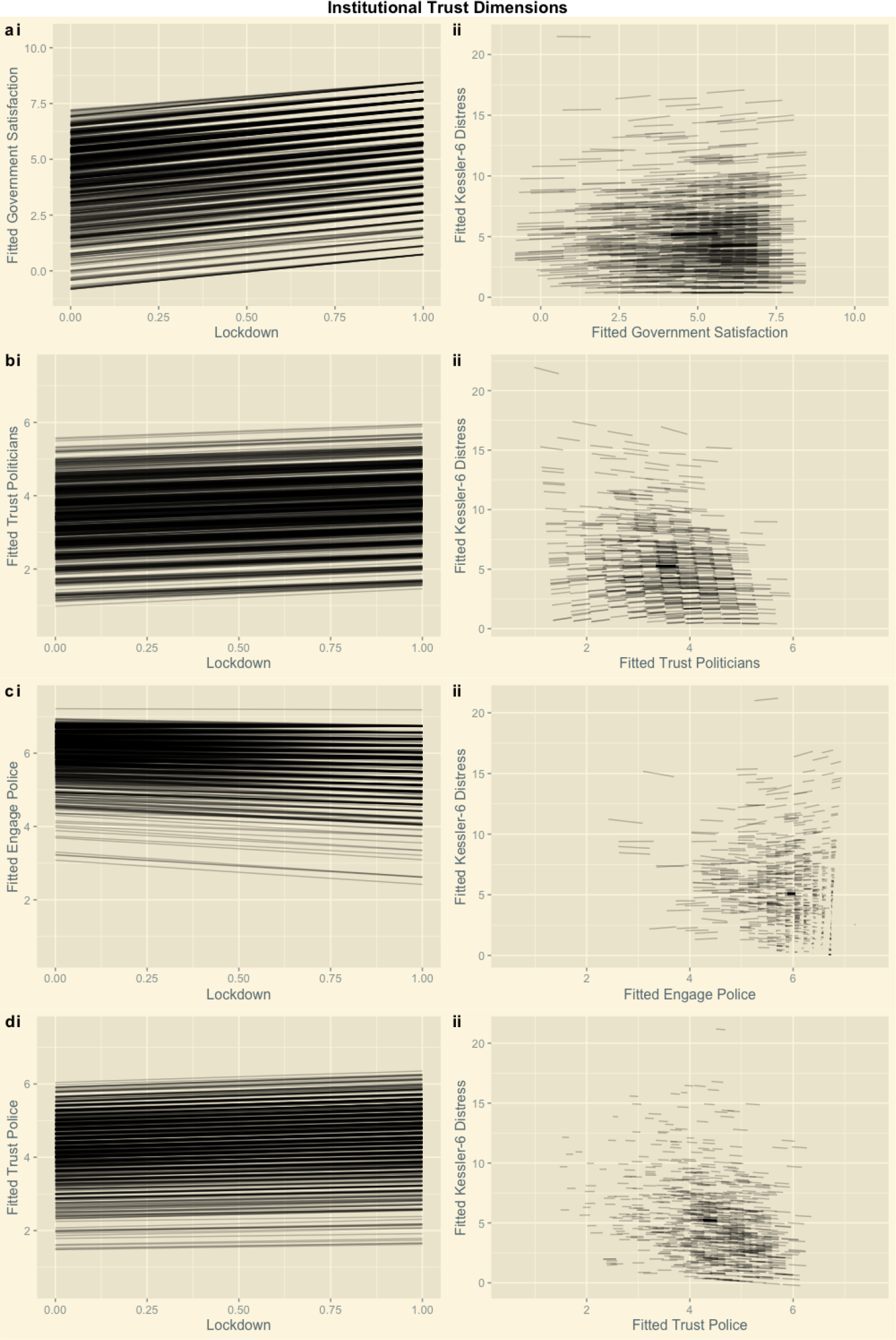
Predictive Plots: Institutional Trust. Panel (i) presents the fitted values for the ‘a’ path X→M. Panel (ii) presents the fitted values for the ‘b’ path (M→Y). We do not find evidence for strong relationships between institutional trust indicators and psychological distress (the b paths).

**Figure B5.**
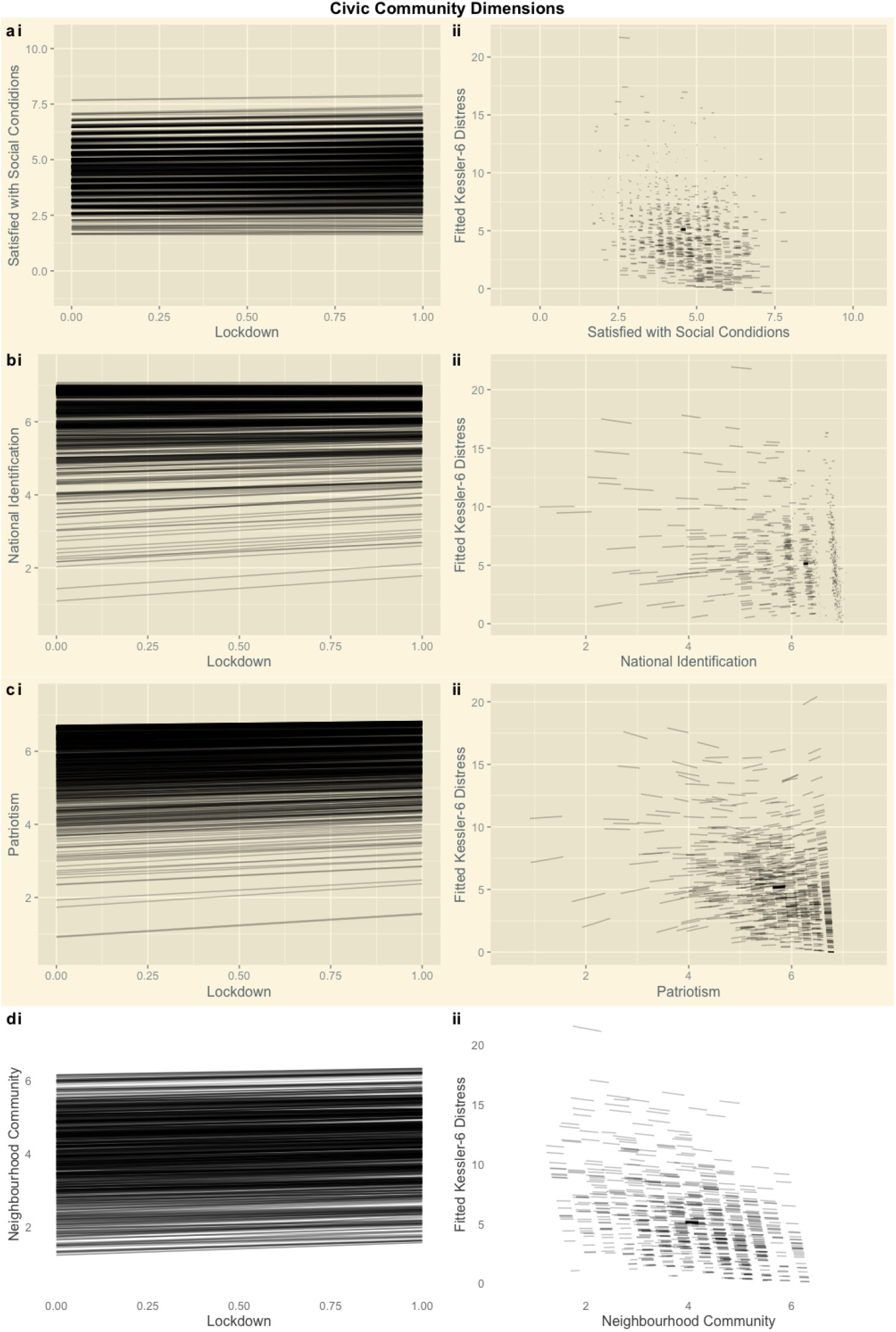
Predictive Plots: Civic Attitudes. Panel (i) presents the fitted values for the ‘a’ path (X→M). Panel (ii) presents the fitted values for the ‘b’ path (M→Y). We do not observe reliable mediation effects for satisfaction with social conditions, national identification, or patriotism. Of these we observe a strong relationship between satisfaction with social conditions and psychological distress (the ‘b’ path), which reveals that if this indicator had been compromised during the lockdown then New Zealanders would have experienced greater psychological distress. The white panels (d.i, d.ii) describe the ‘a’ and ‘b’ paths for the reliable mediation effect of the lockdown on psychological distress through increased neighbourhood community. Specifically, the lockdown increased neighbourhood community, and furthermore, because greater neighbourhood community predicted lower psychological distress, there was a buffering mediation effect.

## Appendix C Background

As New Zealand entered its first national COVID-19 lock-down at midnight on 25^th^ March, the country confronted three pandemic challenges to mental health. First, the lock-down occurred in a setting of nationwide health uncertainty. On 25^th^ February, the New Zealand Ministry of Health indicated that ICU beds across regional DHBs totalled 176 (“COVID-19 (novel coronavirus) update −25 February,” 2020), less than 3.5 beds for every 100,000 residents. On 28^th^ February, the country declared its first COVID-19 case, and on 5^th^ March, the country reported its first case of person-to-person transmission. On 19^th^ March, New Zealand closed its borders to residents and non-citizens. On 20^th^ March, New Zealand announced its 4-level COVID-19 Alert System, which described a series of progressively restrictive state-mandated physical distancing restrictions (“Alert system overview,” 2020). That same day, the country moved to Alert Level 2. On 23^rd^ March, the Ministry of Health reported its first case of suspected community transmission, and that day moved to Alert Level 3; all public gatherings, including funerals and religious worship, were prohibited by the government on the same day. When the country moved into Alert Level 4 (nationwide lockdown), New Zealand’s COVID-19 infection rates were taking off exponentially. New Zealand’s disease modellers predicted that without stringent physical distancing measures, the surge of severe COVID-19 infections would overwhelm the country’s healthcare system by June (James et al., 2020). On 1^st^ April, 61 new cases were reported; On 2^nd^ April, there were 89 new cases; On April 3^rd^, there were 71 new cases; On April 4^th^, there were 82 new cases. By April 12^th^, New Zealand had already reported its second death. Despite high levels of public compliance with restrictive lockdown measures (Google, 2020; “HorizonPoll - Horizon Research NZ online poll,” 2020), a geometric growth in mortality soon followed. Epidemiological models predicted that even a reduction of the virus transmission rate to *R*_0_ = 1.2 would eventually result in the deaths of 1.58% of the country’s population (James et al., 2020). At the same time, images of Wuhan, Lombardy, and New York City were flooding the country.

Second, New Zealand’s severe economic and social March/April 2020 lockdown occurred in a setting of pervasive economic uncertainty. By 3^rd^ February, the New Zealand Government restricted entry for those travelling from mainland China, closing a key tourism market. By early March, local and global financial markets were spiralling downward. On 16^th^ March, the Government halted share trading and the New Zealand Reserve Bank announced emergency cuts to its lending rate. On 17^th^ March, the New Zealand government announced an emergency economic support package for businesses and workers valued at 4% of national GDP (NZD $12.1B)(Strongman, 2020). Financial forecasters were predicting declines in New Zealand’s GDP of at least 5% and at least a doubling of the country’s unemployment rate (“Budget Economic and Fiscal Update 2020,” 2020).

Third, New Zealand’s Alert Level 4 restrictions during March/April 2020 were amongst the most severe in the world, resulting in a sudden loss of ordinary economic and social routines for most of the population. Specifically, New Zealand’s Alert Level 4 lockdown mandated: (1) home-based isolation and limiting of contact to households; (2) restrictions on recreational activity: some forms of isolated recreational activity were permitted, but only in one’s local area; (3) limiting travel to essential activities or services; (4) cancellations of all gatherings, including funerals, and closure of all public venues; (5) closure of all businesses except for essential services (supermarkets, pharmacies, clinics, petrol stations and lifeline utilities); (6) closure of all educational facilities; (7) rationing of supplies and, where needed, the requisitioning of facilities for managed isolation and COVID-19 testing; and (8), the re-prioritisation of healthcare services to manage COVID-19 disease progression (“Alert system overview,” 2020). An independent stringency test ranked New Zealand the 8^th^ most restrictive country, scoring 96.30 out of a possible 100 severity points (“COVID-19: Government Response Stringency Index,” 2020).

Despite suffering the health and economic uncertainties that other countries witnessed early in their COVID19 pandemics, New Zealand’s stringent Alert Level 4 restrictions in March/April 2020 also occurred in a setting of widespread government confidence (Sibley, Greaves, et al., 2020). For example, each day of New Zealand’s COVID19 lockdown, New Zealand’s Ministry of Health, often with New Zealand’s Prime Minister as spokes-person, reassured the nation about the availability of essential supplies, provided transparent updates about COVID-19 case histories, and described details of New Zealand’s pandemic health and economic planning, with explicit clarity about areas of pandemic uncertainty, such as the duration of the lockdown. In the second week of New Zealand’s Alert Level 4, the government launched a dedicated COVID-19 WhatsApp channel for case updates and health information and encouraged residents to share their personal stories using the #BeKind hashtag. That same week, the Ministry of Education announced the development of distance learning infrastructure at all levels of public education in response to student learning and wellbeing needs during the lockdown. The initiative included the delivery of learning materials and digital devices to students, online resources for parents, professional development options for teachers, and the aim to connect more New Zealand residences to the Internet. In addition, initiatives for transitioning from Alert Level 4 to Alert Level 3 were announced by Prime Minister Jacinda Arden on Day 15 of COVID-19 lockdown, providing clarity on how the transition to less restrictive economic and social pandemic regimes would eventually occur.Additionally, although lockdown measures were strict, Google mobility data for March 29*^th^* shows steeply declining mobility trends suggesting New Zealanders were compliant with the measures (Google, 2020), an observation concurrent with poll results from Horizon Research NZ indicating 95% of New Zealand adults reported complying with lockdown measures between April 7*^th^* and 12*^th^* (“HorizonPoll - Horizon Research NZ online poll,” 2020).

Though New Zealand resembled other countries in its confrontation with stark economic and health uncertainties, pandemic lockdown occurred in a context of high institutional confidence, early government pandemic action, and effective policy and science communication. These benefits were lacking in other countries. High public confidence minimises potentially confounding distress caused from dissatisfaction with the government response, enabling greater clarity about specific distress mechanisms during pandemic lockdown.

## Appendix D Extended Method

### Ethics

The New Zealand Attitudes and Values Study was approved by The University of Auckland Human Participants Ethics Committee on 03-June-2015 until 03-June-2018, and renewed on 05-September-2017 until 03-June-2021. Reference Number: 014889. Our previous ethics approval statement for the 2009-2015 period is: The New Zealand Attitudes and Values Study was approved by The University of Auckland Human Participants Ethics Committee on 09-September-2009 until 09-September-2012, and renewed on 17-February-2012 until 09-September-2015. Reference Number: 6171. All participants granted informed written consent and The University of Auckland Human Participants Ethics Committee approved all procedures.

### Participants

#### Demographics

**Age**. The mean age of our sample was 50.7 (*SD* = 13.3) in 2018 and 51.9 (*SD* = 13.3) in 2020.

**Male**. Gender is assessed in the NZAVS by asking participants to respond using an open-ended question: “What is your gender?” Those who responded as “Male” were coded as “1” and “Not Male” were coded as “0.”

**Deprivation**. We measure the socioeconomic status of participants’ immediate (small area) neighborhood using the 2013 New Zealand Deprivation Index, which uses aggregate census information about the residents of small neighborhood-type units to assign a decile-rank index from 1 (least deprived) to 10 (most deprived) (Atkinson et al., 2014). The index is based on a Principal Components Analysis of the following nine variables (in weighted order): proportion of adults who received a means-tested benefit, household income, proportion not owning own home, proportion single-parent families, proportion unemployed, proportion lacking qualifications, proportion household crowding, proportion no telephone access, and proportion no car access. Thus, the index reflects the average level of deprivation for small neighborhood-type units (or small community areas of approximately 80–90 people each) across the entire country.

**Education**. Education level was measured using an 11-point rating developed by the New Zealand government known as the New Zealand Qualification Framework (NZQF; 0 = no qualification, 10 = doctoral degree). Sample demographic statistics are reported in TableD1.

**European**. Ethnicity was assessed using two basic categories: (1) New Zealand European/Pākehā coded as “1” and (2) others coded as “0.”

**Religious**. To assess religiousness, we asked people: “Do you identify with a religion and/or spiritual group?” (yes or no). We coded “yes” as “1” and “no” as “0.”

We report sample descriptive statistics for sample demographic statistics in D1.

**Table D1.** Demographic parameters

|  | Wave |  |
| --- | --- | --- |
|  | 2018<br>n = 940 | 2020L4<br>n = 940 |
| Age | 50.7 (13.3) | 51.9 (13.3) |
| Male |  |  |
| Not Male | 611 (65%) | 614 (65.3%) |
| Male | 326 (34.7%) | 326 (34.7%) |
| missing | 3 (0.3%) | 0 (0%) |
| European Ethnicity |  |  |
| Not European | 72 (7.7%) | 57 (6.1%) |
| European | 868 (92.3%) | 883 (93.9%) |
| missing | 0 (0%) | 0 (0%) |
| Education | 5.7 (2.6) | 5.6 (2.7) |
| NZ Deprivation | 4.6 (2.7) | 4.6 (2.7) |
| Religious |  |  |
| Not_Religious | 607 (64.6%) | 628 (66.8%) |
| Religious | 327 (34.8%) | 312 (33.2%) |
| missing | 6 (0.6%) | 0 (0%) |

### Predictors of Psychological Distress

#### Health Concerns, Rumination, and Fatigue

Following Sibley, Greaves, et al. (2020)’s preregistered study, we investigated both subjective health responses as well as the frequency of health states associated with psychological distress: rumination and feelings of exhaustion (fatigue).

**Fatigue**. We measure participants’ subjective fatigue by asking, “During the last 30 days, how often did…you feel exhausted?”. Responses to this item were rated on an ordinal scale ranging from none of the time (0) to all of the time (4).

**Rumination**. We measure rumination using an item adapted from Nolen-Hoeksema and Morrow (1993), “During the last 30 days, how often did…you have negative thoughts that repeated over and over?”. Responses to this item were rated on an ordinal scale ranging from none of the time (0) to all of the time (4).

**Satisfaction with Health**. The link between health satisfaction and psychological distress is firmly established (Cockerham et al., 1988; Han et al., 2015; G. Williams et al., 2017). A recent study identified fear of COVID-19 as a reliable predictor of positive behavioral change and compliance with public health measures (Harper et al., 2020). Another study noted that public perception of health risks plays a key role in the adoption of mitigation measures (Motta Zanin et al., 2020). We measure health satisfaction using an item taken from the Australian Unity Wellbeing Index (Cummins et al., 2017). Participants rated their satisfaction with “Your health” on a 10-point Likert scale ranging from completely dissatisfied (1) to completely satisfied (10).

**Short-Form Health**. We measure subjective health using three items taken from the MOS 36-item short-form health survey (Ware Jr & Sherbourne, 1992):

1. “In general, would you say your health is…”;
2. “I seem to get sick a little easier than most people”;
3. “I expect my health to get worse.”

Responses to these items were rated on a 7-point Likert scale ranging from poor (1) to excellent (7) (reverse coded sub-scales 2, 3).

We report sample descriptive statistics for indicators of health in D2.

#### Economic Concerns

**Future Security**. We measure satisfaction with future security using an item taken from the Australian Unity Wellbeing Index (Cummins et al., 2017). Participants rated their level of satisfaction with “Your future security” on a 10point Likert scale ranging from completely dissatisfied (1) to completely satisfied (10).

**Satisfaction with Business**. Previous research finds that economic concerns affect distress (McKee-Ryan et al., 2005), which may play a role in COVID-19 distress dynamics (Collie et al., 2020). We measure business satisfaction using an item taken from the National Wellbeing Index (Tiliouine et al., 2006). Participants rated their level of satisfaction with “Business in New Zealand” on a 10-point Likert scale ranging from completely dissatisfied (1) to completely satisfied (10).

**Table D2.** Indicators of Health

|  | Wave |  | P-Value |
| --- | --- | --- | --- |
|  | 2018<br>n = 940 | 2020L4<br>n = 940 |  |
| Fatigue |  |  | <.001 |
| None Of The Time | 118 (12.6%) | 164 (17.4%) |  |
| A Little Of The Time | 299 (31.8%) | 320 (34%) |  |
| Some Of The Time | 293 (31.2%) | 293 (31.2%) |  |
| Most Of The Time | 165 (17.6%) | 128 (13.6%) |  |
| All Of The Time | 58 (6.2%) | 34 (3.6%) |  |
| missing | 7 (0.7%) | 1 (0.1%) |  |
| Rumination |  |  | 0.242 |
| None Of The Time | 469 (49.9%) | 465 (49.5%) |  |
| A Little Of The Time | 248 (26.4%) | 280 (29.8%) |  |
| Some Of The Time | 151 (16.1%) | 148 (15.7%) |  |
| Most Of The Time | 50 (5.3%) | 33 (3.5%) |  |
| All Of The Time | 14 (1.5%) | 13 (1.4%) |  |
| missing | 8 (0.9%) | 1 (0.1%) |  |
| Satisfied with Health | 6.6 (2.4) | 6.5 (2.4) | 0.525 |
| Short Form Health | 4.9 (1.2) | 5.0 (1.2) | 0.471 |

**Table D3.**
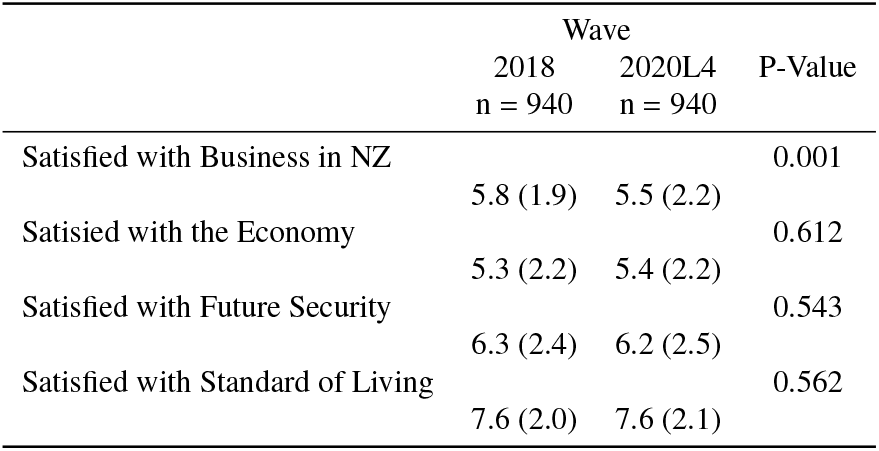
Indicators of Economic Security

**Satisfaction with the Economy**. We measure satisfaction with the New Zealand economy using an item taken from the National Wellbeing Index (Tiliouine et al., 2006). Participants evaluated “The economic situation in New Zealand” on a 10-point Likert scale ranging from completely dissatisfied (1) to completely satisfied (10).

**Satisfaction with Standard Of Living**. We assess satisfaction with standard of living using an item taken from the Australian Unity Wellbeing Index (Cummins et al., 2017). Participants rated their level of satisfaction with “Your standard of living” on a 10-point Likert scale ranging from completely dissatisfied (1) to completely satisfied (10).

We report sample descriptive statistics for indicators of economic attitude in D2.

#### Personal Relationships

**Social Support**. We measure perceived social support using a three items taken from Cutrona and Russell (1987) and K. D. Williams et al. (2000).

1. “There are people I can depend on to help me if I really need it”;
2. “There is no one I can turn to for guidance in times of stress”;
3. “I know there are people I can turn to when I need help.”

Responses to this item were rated on a 7-point Likert scale ranging from strongly disagree (1) to strongly agree (7) (reverse coded subscale 2).

**Table D4.** Personal Relationships

|  | Wave |  | P-Value |
| --- | --- | --- | --- |
|  | 2018<br>n = 940 | 2020L4<br>n = 940 |  |
| Partner |  |  | 0.998 |
| 0 | 229 (24.4%) | 236 (25.1%) |  |
| 1 | 679 (72.2%) | 704 (74.9%) |  |
| missing | 32 (3.4%) | 0 (0%) |  |
| Satisfied with Personal Relationships | 7.7 (2.1) | 6.5 (2.4) | <.001 |
| Social Support | 6.0 (1.1) | 6.0 (1.2) | 0.336 |
| Social Belonging | 5.1 (1.1) | 5.1 (1.1) | 0.569 |

**Social Belonging**. Previous research finds a strong relationship between social belonging and distress (Haslam et al., 2012; Holt-Lunstad et al., 2015). Belonging, or the lack thereof, is a direct predictor of depression (Choenarom et al., 2005; Hagerty & Williams, 1999). We measure social belonging using three items adapted from the Sense of Belonging Instrument (Hagerty & Patusky, 1995):

1. “Know that people in my life accept and value me”;
2. “Feel like an outsider”;
3. “Know that people around me share my attitudes and beliefs.”

Responses to this item were rated on a 7-point Likert scale ranging from very inaccurate (1) to very accurate (7) (reverse coded subscale 2). This indicator was centered and standardised.

**Satisfaction with Personal Relationships**. We measure satisfaction with personal relationships using an item taken from the Australian Unity Wellbeing Index (Cummins et al., 2017). Participants rated their satisfaction with “Your personal relationships” on a 10-point Likert scale ranging from completely dissatisfied (1) to completely satisfied (10).

We report sample descriptive statistics for indicators of personal relationships in D4.

#### Institutional Trust

**Satisfaction with the Government**. In New Zealand, government confidence rose markedly during COVID-19 lockdown and Sibley, Greaves, et al. (2020) speculate that confidence in government helped to mitigate COVID-19 related distress. Here, we assess government confidence using an item adapted from the National Wellbeing Index (Tiliouine et al., 2006). Participants evaluated “The performance of the current New Zealand Government” on a 10-point Likert scale ranging from completely dissatisfied (1) to completely satisfied (10). This indicator was centered and standardised.

**Trust in Politicians**. We measure trust in politicians by asking participants to rate their agreement with the statement: “Politicians in New Zealand can generally be trusted”. Responses to this item were rated on a 7-point Likert scale ranging from strongly disagree (1) to strongly agree (7).

**Engagement with Police**. We measure engagement with police using two items taken from Tyler (2005).

1. “I would always report dangerous or suspicious activities occurring in my neighbourhood to the police”;
2. “I would always provide information to the police to help them find someone suspected of committing a crime.”

Responses to these items were rated on a 7-point Likert scale ranging from strongly disagree (1) to strongly agree (7).

**Trust in Police**. We measure institutional trust in police using three items taken from Tyler (2005).

1. “People’s basic rights are well protected by the New Zealand Police”;
2. “There are many things about the New Zealand Police and its policies that need to be changed”;
3. “The New Zealand Police care about the well-being of everyone they deal with.”

Responses to these items were rated on a 7-point Likert scale ranging from strongly disagree (1) to strongly agree (7) (reverse coded subscale 2).

We report sample descriptive statistics for indicators of institutional trust in D5.

#### Civic Community

**Satisfaction with Social Conditions**. We measure satisfaction with social conditions in New Zealand using an item taken from the National Wellbeing Index (Tiliouine et al., 2006). Participants rated their level of satisfaction with “The social conditions in New Zealand” on a 10-point Likert scale ranging from completely dissatisfied (1) to completely satisfied (10).

**Table D5.** Indicators of Institutional Trust

|  | Wave |  | P-Value |
| --- | --- | --- | --- |
|  | 2018<br>n = 940 | 2020L4<br>n = 940 |  |
| Satisfied with Government | 5.6 (2.6) | 7.1 (2.5) | <.001 |
| Trust Politicians | 3.7 (1.4) | 4.1 (1.4) | <.001 |
| Trust Police | 4.5 (1.2) | 4.8 (1.2) | <.001 |
| Engage with Police | 5.9 (1.1) | 5.7 (1.2) | 0.016 |

**National Identity**. We measure identification with New Zealand using a single-item measure of social identification taken from Postmes et al. (2013). Participants evaluated “I identify with New Zealand” on a 7-point Likert scale ranging from strongly disagree (1) to strongly agree (7).

**Patriotism**. We measure patriotism using two items adapted from Kosterman and Feshbach (1989).

1. “I feel a great pride in the land that is our New Zealand”;
2. “Although at time I may not agree with the government, my commitment to New Zealand always remains strong.”

Responses to these items were rated on a 7-point Likert scale ranging from strongly disagree (1) to strongly agree (7).

**Sense of Neighbourhood Community**. We postulate that community sensibilities may buffer mental distress during mass crises (Sibley, Greaves, et al., 2020, see also: Drury, 2012). We measure a sense of community by asking participants to rate their agreement with the statement: “I feel a sense of community with others in my local neighbourhood.” Responses to this item were rated on a 7-point Likert scale ranging from strongly disagree (1) to strongly agree (7). This indicator was centered and standardised.

We report sample descriptive statistics for indicators of civic community in D6.

**Table D6.**
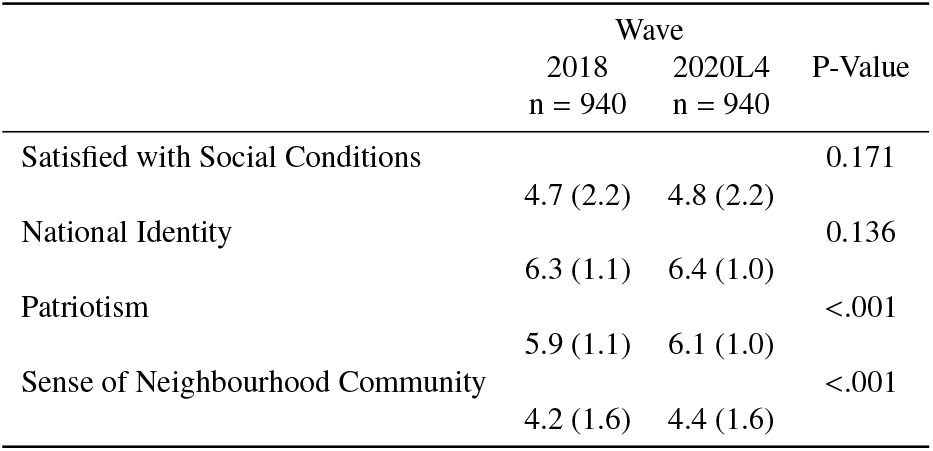
Indicators of Civic Community

|  | Wave |  | P-Value |
| --- | --- | --- | --- |
|  | 2018<br>n = 940 | 2020L4<br>n = 940 |  |
| Satisfied with Social Conditions | 4.7 (2.2) | 4.8 (2.2) | 0.171 |
| National Identity | 6.3 (1.1) | 6.4 (1.0) | 0.136 |
| Patriotism | 5.9 (1.1) | 6.1 (1.0) | <.001 |
| Sense of Neighbourhood Community | 4.2 (1.6) | 4.4 (1.6) | <.001 |

### Indicator of Psychological Distress: The Kessler-6

We measure psychological distress using the Kessler-6 scale (Kessler et al., 2002), which exhibits strong diagnostic concordance for moderate and severe psychological distress in large, cross-cultural samples (Kessler et al., 2010; Prochaska et al., 2012). Participants rated during the past 30 days, how often did… (a) “... you feel hopeless”; (b) “... you feel so depressed that nothing could cheer you up”; (c) “... you feel restless or fidgety”; (d)”... you feel that everything was an effort”; (e) “... you feel worthless”; (f) “... you feel nervous?” Ordinal response alternatives for the Kessler-6 are: “None of the time”; “A little of the time”; “Some of the time”; “Most of the time”; “All of the time.”

We report sample descriptive statistics for indicators of personal Kessler-6 distress in D7.

**Table D7.** Indicators of Civic Community

|  | Wave |  | P-Value |
| --- | --- | --- | --- |
|  | 2018<br>n = 940 | 2020L4<br>n = 940 |  |
| K6 (summed) | 5.4 (4.0) | 5.7 (3.8) | 0.034 |
| K6 diagnostic categories |  |  | 0.366 |
| Low Distress | 539 (57.3%) | 515 (54.8%) |  |
| Moderate Distress | 337 (35.9%) | 369 (39.3%) |  |
| Severe Distress | 58 (6.2%) | 56 (6%) |  |
| missing | 6 (0.6%) | 0 (0%) |  |

### Missingness

99.6% of the dataset was complete. The columns with the highest rates of missingness were willingness to engage the police (2.02% missing), trust in politicians (.96% missing), identify with New Zealand (1.17%), satisfied with the New Zealand economy (.8% missing), and satisfied with business conditions in New Zelaand (.48%) missing. Though rates of overall missingness were low, to avoid biasing estimates, we multiply imputed missing values using the Amelia package in R(Honaker et al., 2011). We passed the individual id column indicator to Amelia a the clustering indicator, and other columns in the dataset were multiply imputed under a missing at random (MAR) assumption (i.e. random conditional on the information contained within all variables in the dataset).

### Statistical models

We fit a series of Bayesian multilevel mediation models using the bmlm package in RVuorre, 2017. Estimation of direct and mediation (or “indirect”) effects in a Bayesian setting is straightforward (Yuan & MacKinnon, 2009). We simultaneously fit an outcome model in which K6 summed scores were regressed on the time interval (pre/during lockdown, the direct effect) and the mediator, and a mediator model in which the mediator was regressed on the time interval (direct effect), and both models adjusted for the repeated measures within individuals. The model equations and priors for these models are presented below.

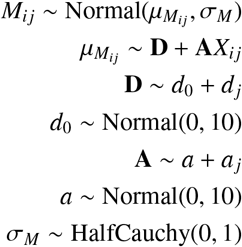

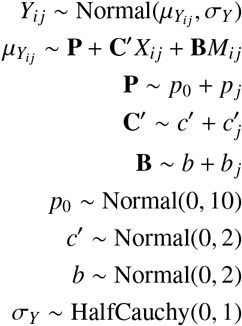

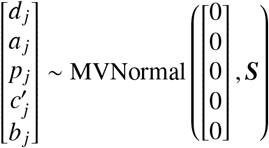

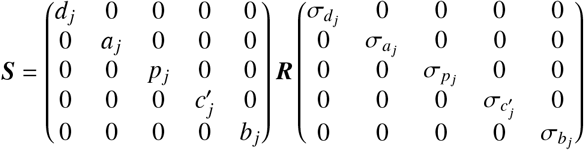

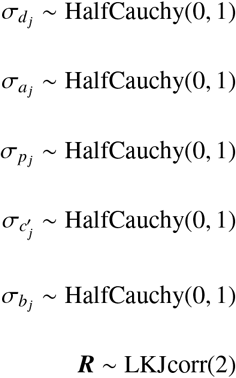

These priors were only weakly informative; we also used the default bmlm priors for the three models in which mediation effects were detected. Minor differences were within the region of MCMC error, with no difference to inferences in this article. We infer that all the information in these models came from the data with no influence from the priors.

In Bayesian estimation, the direct effect, ‘c′’, is the mean value of posterior samples from treatment on the outcome (X→Y), the mediator effect, ‘b’, is the mean value of posterior samples from the mediator on the outcome (X→Y); the meditation effect (or the indirect effect) is the mean value of the multiplication of the posterior samples from the mediator of the outcome model and the posterior samples from treatment of the mediation model (a * b) plus the individual level covariances of these slopes 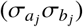. Hence, the mediation effect (or indirect effect) is the product of the posterior distributions for the slope of the direct effect (lockdown) on the mediator, the ‘a’ path (X→M), and the slope of the mediator effect on the outcome (psychological distress), the ‘b’ path (M→Y), plus the covariance of 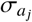 and 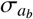, which are the individual level variances of the a and b paths:

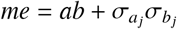

The total effect is:

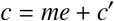

These equations were obtained from Vuorre and Bolger (2018).

Given the size of our sample, we use 95% Credible Intervals for describing uncertainty rather than the default 89% credible intervals that McElreath (2018) advises (among other primes), to make probabilistic reasoning explicit (Makowski, Ben-Shachar, et al., 2019).

We performed statistical analysis in R version 4.0.2 (2020-06-22), Platform: x86-64-apple-darwin17.0 (64-bit), Running under: macOS Catalina 10.15.3.(R Core Team, 2020). We are grateful to authors and contributors of the following R packages that we used in this study (Barrett & Brignone, 2017; Brilleman et al., 2018; Bürkner, 2018; Dahl et al., 2019; Fernández-i-Marín, 2016; Gelman & Su, 2020; Goodrich et al., 2020; Kassambara, 2020; Leifeld, 2013; Lüdecke, 2019; Lüdecke, 2020; Lüdecke, Ben-Shachar, & Makowski, 2020; Lüdecke, Ben-Shachar, Waggoner, et al., 2020; Lüdecke, Makowski, et al., 2020; Makowski, Ben-Shachar, et al., 2019; Rich, 2020; Wickham, 2016; Wickham et al., 2019). We’re especially grateful for Daniel Lüedecke and Matti Vourre for general advice, though any errors remain our responsibility.

## Appendix E Sampling Procedure

The NZAVS is a national longitudinal panel study that has been tracking social attitudes, personality, and health outcomes within individuals since 2009 (*Ns* = 6,441-47,951). Here, we conduct a novel analysis of the rolling 2020 data-frame first used by Sibley, Greaves, et al. (2020). The longitudinal sample is composed of 940 participants who responded to both Wave 10 (2018) and Wave 11 (2020) of the NZAVS, which were taken respectively before and during the early stages of New Zealand’s first COVID-19 lockdown in March/April 2020. The Time 10 (2018) NZAVS contained responses from 47,951 participants (18,010 retained from one or more previous waves). The sample retained 2,964 participants from the Time 1 (2009) sample (a retention rate of 45.5%). The sample retained 14,049 participants from Time 9 (2017; a retention rate of 82.3% from the previous year). Participants who provided an email address were first emailed and invited to complete an online version if they preferred. Participants who did not complete the online version (or did not provide an email) were then posted a copy of the questionnaire, with a second postal follow-up two months later. We staggered the time of contact, so that participants who had completed the previous wave were contacted approximately one year after they last completed the questionnaire. We offered a prize draw for participation (five draws each for $1000 grocery vouchers, a prize pool total of $5000). All participants were posted a Season’s Greetings card from the NZAVS research team and informed that they had been automatically entered into a bonus seasonal grocery voucher prize draw. Participants were also emailed an eight-page newsletter about the study.

To boost sample size and increase sample diversity for subsequent waves, a booster sample was conducted by selecting people from the New Zealand electoral roll. As with previous booster samples, sampling was conducted without replacement (i.e., people included in previous sample frames were identified and removed from the 2018 roll). Booster samples completed over the first decade of the study mean that the NZAVS has been drawn from random electoral roll samples, stratified electoral roll samples, and random electoral roll samples with upper age limits. Wave-on-wave retention has generally been high, usually upwards of 80%. As such, all the participants analysed in this paper have completed at least one other survey, providing assurance that there is nothing about the current sample in terms of sudden opting-in during the pandemic or lockdown. A detailed description of the sampling procedure for the NZAVS is available in Sibley, Greaves, et al. (2020).

The NZAVS 2018 sample frame consisted of 325,000 people aged from 18-65 randomly selected from the 2018 New Zealand Electoral Roll, who were currently residing in New Zealand (one can be registered to vote in New Zealand but living overseas). The electoral roll contained 3,250,000 registered voters. The New Zealand Electoral Roll contains participants’ date of birth (within a one-year window), and we limited our frame to people who are 65 or younger, due to our aim of retaining participants longitudinally. We concurrently advertised the survey on Facebook via a $5000 paid promotion of a link to a YouTube video describing the NZAVS and the large booster sample we were conducting. The advertisement ran for 14 days and targeted men and women aged 18-65+ who lived in New Zealand. This paid promotion reached 147,296 people, with 4,721 link clicks (i.e., clicking to watch the video), according to Facebook. The goal of the paid promotion was twofold: (a) to increase name recognition of the NZAVS during the period in which questionnaires were being posted, and (b) to help improve retention by potentially reaching previous participants who happened to see the advertisement. A total of 29,293 participants who were contained in our sample frame completed the questionnaire (response rate = 9.2% when adjusting for the 98.2% accuracy of the 2018 electoral roll). A further 648 participants completed the questionnaire, but were unable to be matched to our sample frame (for example, due to a lack of contact information) or were unsolicited opt-ins. Informal analysis indicates that unsolicited opt-ins were often the partners of existing participants.

The longitudinal data used in the study were collected for a complementary analysis in Sibley, Greaves, et al. (2020) to provide a within-person comparison for the pre-registered propensity-score matched analysis. In Sibley, Greaves, et al. (2020) we report the results of paired sample t-tests for the within-subjects comparisons and find these largely match the separate propensity-score matched analysis. Specifically, we observe: “Sense of community increased (*p* <.001, Cohen’s d = .17), as did mental distress (*p* = .027, Cohen’s d = .06).” Apart from this paired sample t-test of average population-level distress before and during lockdown in our follow-up within-subjects analyses, we do not conduct any further analysis of psychological distress using the panel sample in Sibley, Greaves, et al. (2020).

### Sample Comparison

To assess systematic biases in the online-only New Zealand Attitudes and Values responses, we compared the lockdown sample at baselines in 2018/19 with the full 2018/2019 sample baselines. These comparisons are reported in Tables E1, E2, E4, E5, and E6.

Compared with the full 2018 sample, in 2018 the lockdown sample was slightly older (M*_fullsample_*_:2018_ = 48.5 (13.9), M_(_*_pre_*_)_*_lockdown_*_:2018_ = 50.7 (13.5), *p* <.001), more educated (M*_fullsample_*_:2018_ = 5.3 (2.7), M_(_*_pre_*_)_*_lockdown_*_:2018_ = 5.7 (2.6), *p* <.001), more European (*_fullsample_*_:2018_ = 88.5%, _(_*_pre_*_)_*_lockdown_*_:2018_ = 92.3%), had slightly worse health as measured by the Short Form Health scale M*_fullsample_*_:2018_ = 5.0 (1.2), M_(_*_pre_*_)_*_lockdown_*_:2018_ = 4.9 (1.2), *p* <.001), and were slightly more trusting of the police M*_fullsample_*_:2018_ = 4.4(1.2), M_(_*_pre_*_)_*_lockdown_*_:2018_ = 4.5 (1.2), *p* <.001). In other respects the lockdown panel sample at the pre-COVID-19 baseline was not reliably different from the full 2018/19 NZAVS panel.

**Table E1.** 2018/2019 Sample Comparison: Demographic Indicators

|  | Select2020 |  | P-Value |
| --- | --- | --- | --- |
|  | 2018only<br>n = 47011 | 2018lockdown<br>n = 940 |  |
| Age | 48.5 (13.9) | 50.7 (13.3) | <.001 |
| Edu | 5.3 (2.7) | 5.7 (2.6) | <.001 |
| Euro |  |  | <.001 |
| 0 | 5384 (11.5%) | 72 (7.7%) |  |
| 1 | 41627 (88.5%) | 868 (92.3%) |  |
| missing | 0 (0%) | 0 (0%) |  |
| Male |  |  | 0.126 |
| 0 | 29410 (62.6%) | 611 (65%) |  |
| 1 | 17485 (37.2%) | 326 (34.7%) |  |
| missing | 116 (0.2%) | 3 (0.3%) |  |
| NZDep | 4.6 (2.7) | 4.6 (2.7) | 0.836 |
| Religious |  |  | 0.415 |
| 0 | 29017 (61.7%) | 607 (64.6%) |  |
| 1 | 16580 (35.3%) | 327 (34.8%) |  |
| missing | 1414 (3%) | 6 (0.6%) |  |

**Table E2.** 2018/2019 Sample Comparison: Health Indicators

|  | Select2020 |  | P-Value |
| --- | --- | --- | --- |
|  | 2018only<br>n = 47011 | 2018lockdown<br>n = 940 |  |
| Satisfied with Health | 6.8 (2.3) | 6.6 (2.4) | 0.035 |
| Short Form Health | 5.0 (1.2) | 4.9 (1.2) | 0.002 |
| Fatigue |  |  | 0.08 |
| None Of The Time | 7209 (15.3%) | 118 (12.6%) |  |
| A Little Of The Time | 14945 (31.8%) | 299 (31.8%) |  |
| Some Of The Time | 14510 (30.9%) | 293 (31.2%) |  |
| Most Of The Time | 7283 (15.5%) | 165 (17.6%) |  |
| All Of The Time | 2514 (5.3%) | 58 (6.2%) |  |
| missing | 550 (1.2%) | 7 (0.7%) |  |
| Rumination |  |  | 0.71 |
| None Of The Time | 22495 (47.9%) | 469 (49.9%) |  |
| A Little Of The Time | 12877 (27.4%) | 248 (26.4%) |  |
| Some Of The Time | 7448 (15.8%) | 151 (16.1%) |  |
| Most Of The Time | 2750 (5.8%) | 50 (5.3%) |  |
| All Of The Time | 854 (1.8%) | 14 (1.5%) |  |
| missing | 587 (1.2%) | 8 (0.9%) |  |

**Table E3.** 2018/2019 Sample Comparison: Economic Indicators

|  | Select2020 |  | P-Value |
| --- | --- | --- | --- |
|  | 2018only<br>n = 47011 | 2018lockdown<br>n = 940 |  |
| Satisfied with Business | 5.8 (1.9) | 5.8 (1.9) | 0.617 |
| Satisfied with the Economy | 5.4 (2.2) | 5.3 (2.2) | 0.195 |
| Satisfied with Future Security | 6.3 (2.3) | 6.3 (2.4) | 0.383 |
| Satisfied with Standard of Living | 7.6 (2.0) | 7.6 (2.0) | 0.412 |

**Table E4.** 2018/2019 Sample Comparison: Personal Relationships Indicators

|  | Select2020 |  | P-Value |
| --- | --- | --- | --- |
|  | 2018only<br>n = 47011 | 2018lockdown<br>n = 940 |  |
| Partner |  |  | 0.978 |
| 0 | 11363 (24.2%) | 229 (24.4%) |  |
| 1 | 33520 (71.3%) | 679 (72.2%) |  |
| missing | 2128 (4.5%) | 32 (3.4%) |  |
| Satisfied with Personal Relationships | 7.7 (2.2) | 7.7 (2.1) | 0.204 |
| Social Belonging | 5.1 (1.1) | 5.1 (1.1) | 0.435 |
| Social Support | 5.9 (1.1) | 6.0 (1.1) | 0.767 |

**Table E5.** 2018/2019 Sample Comparison: Institutional Trust Indicators

|  | Select2020 |  | P-Value |
| --- | --- | --- | --- |
|  | 2018only<br>n = 47011 | 2018lockdown<br>n = 940 |  |
| Satisfied with Government | 5.6 (2.5) | 5.6 (2.6) | 0.188 |
| Trust Politicians | 3.8 (1.4) | 3.7 (1.4) | 0.286 |
| Engage with Police | 5.8 (1.1) | 5.9 (1.1) | 0.231 |
| Trust Police | 4.4 (1.2) | 4.5 (1.2) | 0.02 |

**Table E6.** 2018/2019 Sample Comparison: Civic Community Indicators

|  | Select2020 |  | P-Value |
| --- | --- | --- | --- |
|  | 2018only<br>n = 47011 | 2018lockdown<br>n = 940 |  |
| Patriotism | 5.9 (1.1) | 5.9 (1.1) | 0.66 |
| National Identity | 6.3 (1.1) | 6.3 (1.1) | 0.568 |
| Satisfied with Social Conditions | 4.7 (2.2) | 4.7 (2.2) | 0.461 |
| Sense of Neighbourhood Community | 4.2 (1.7) | 4.2 (1.6) | 0.9 |

**Table E7.**
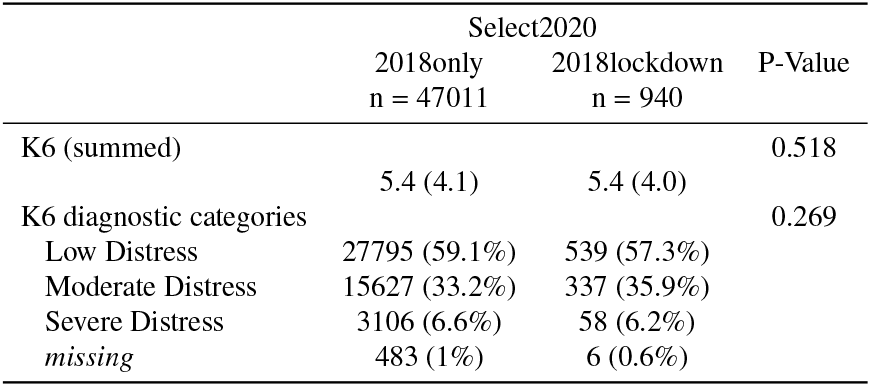
2018/2019 Sample Comparison: Kessler-6 Distress

## Appendix F Mediation Graphs

**Figure F1.**
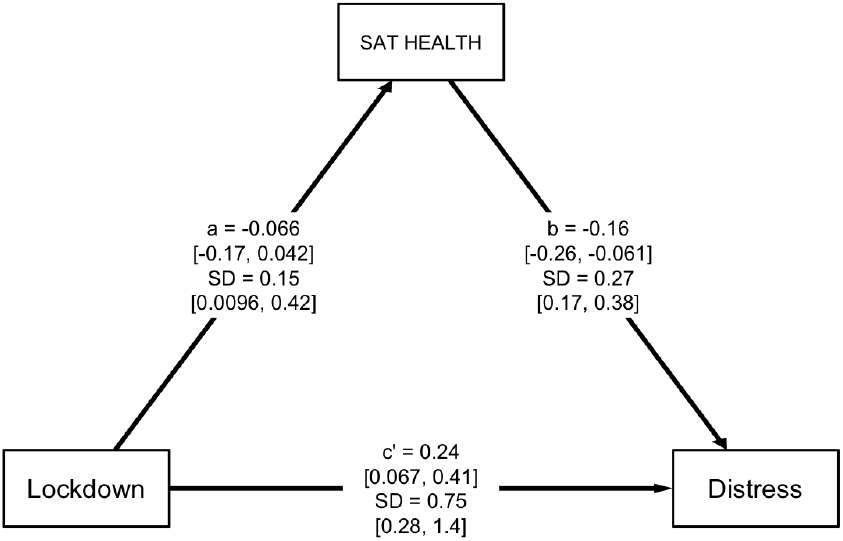
At the population-level average, the lockdown did not reduce satisfaction with health; had it done so, psychological distress would have been worse.

**Figure F2.**
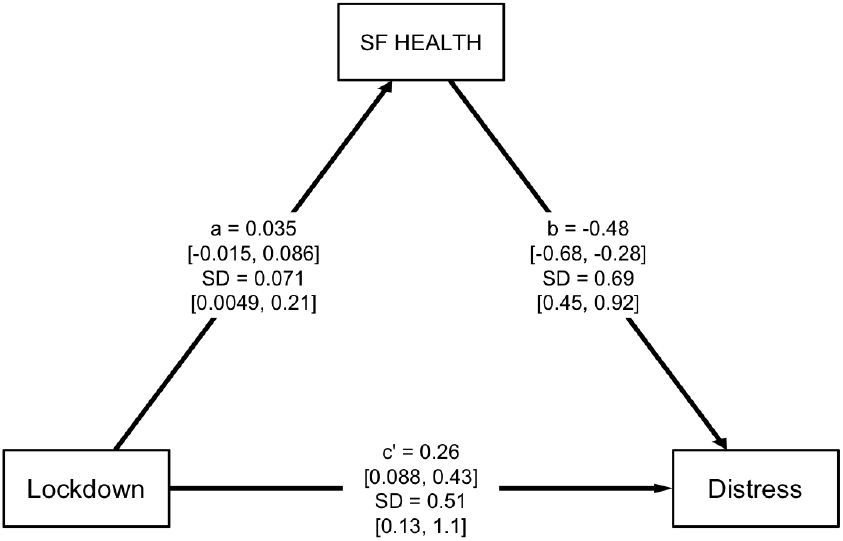
At the population-level average, the lockdown did not reduce short form health scores (i.e., subjective health); had it done so, psychological distress would have been worse.

**Figure F3.**
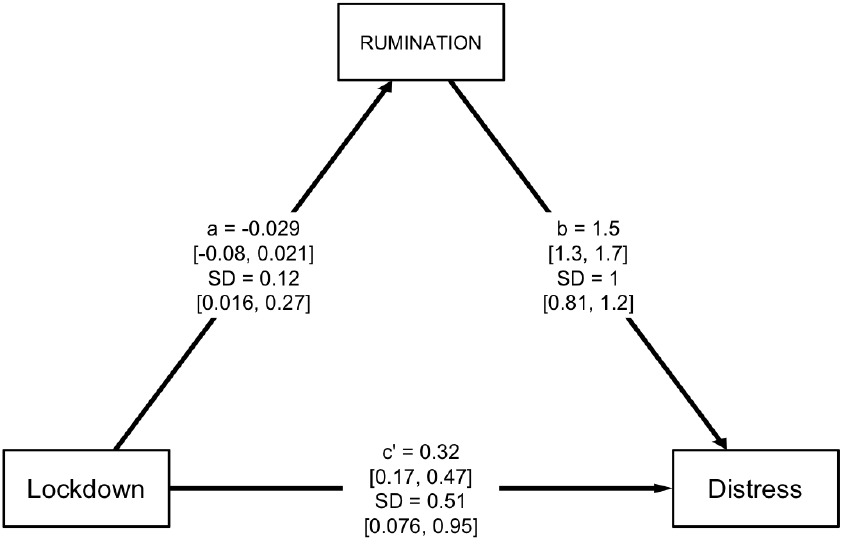
At the population-level average, the lockdown did not increase rumination; had it done so, psychological distress would have been worse.

**Figure F4.**
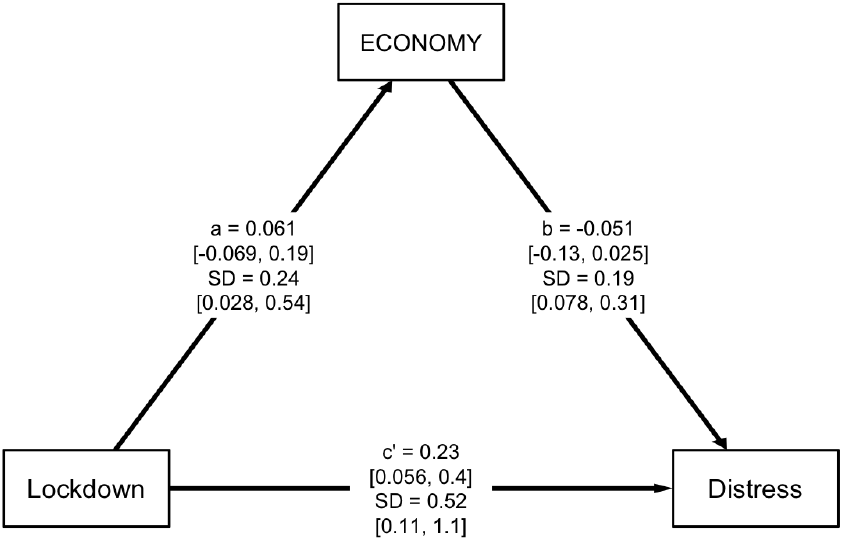
At the population-level average, the lockdown reduced fatigue; moreover, such fatigue reduction buffered people from greater psychological distress.

**Figure F5.**
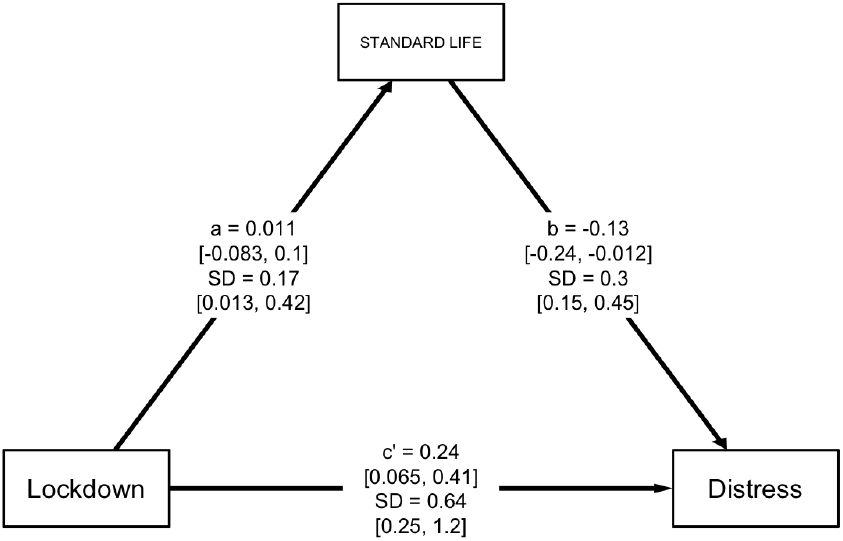
At the population-level average, the lockdown did not reliably affect satisfaction with the economy, and satisfaction with the economy did not reliably predict psychological distress.

**Figure F6.**
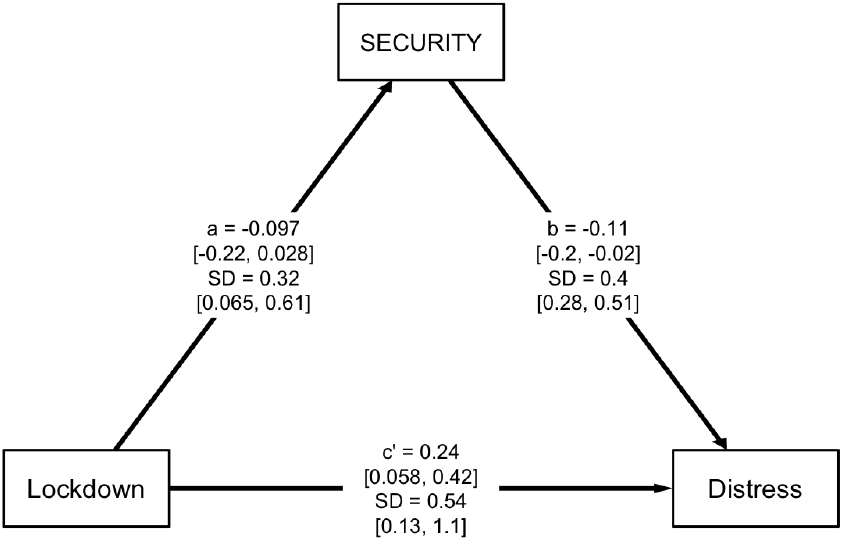
At the population-level average, the lockdown did not reliably affect satisfaction with standard of living; had it done so, psychological distress would have been worse.

**Figure F7.**
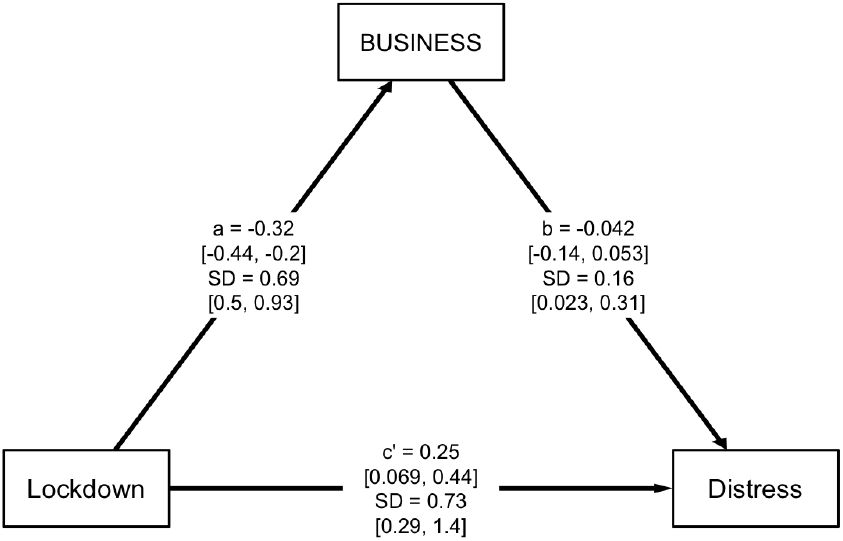
At the population-level average, the lockdown did not reliably affect satisfaction with future security; had it done so, psychological distress would have been worse. Individual-level variation reveals elevated distress in a sub-population who experienced loss of future security.

**Figure F8.**
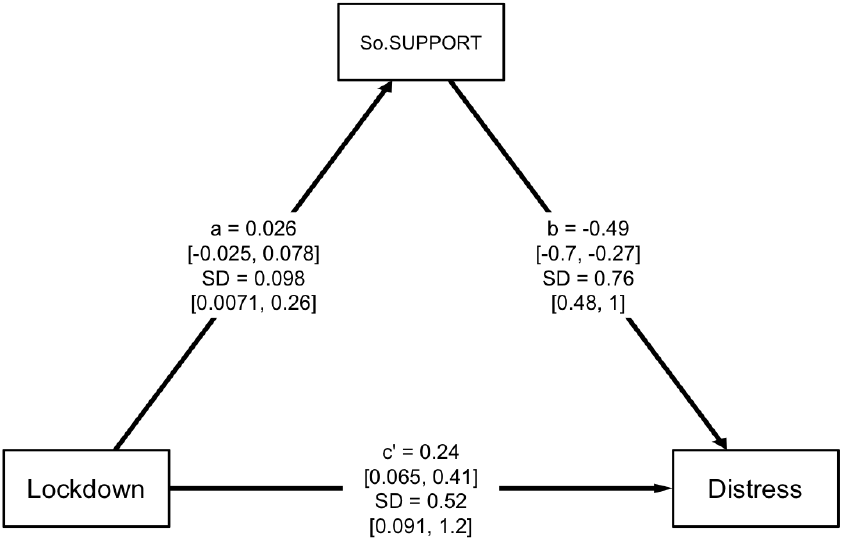
At the population-level average, the lockdown reliably decreased business satisfaction; however, business satisfaction did not reliably predict psychological distress. Nevertheless, there was substantial individual-level variability in the relationship between lockdown and business satisfaction.

**Figure F9.**
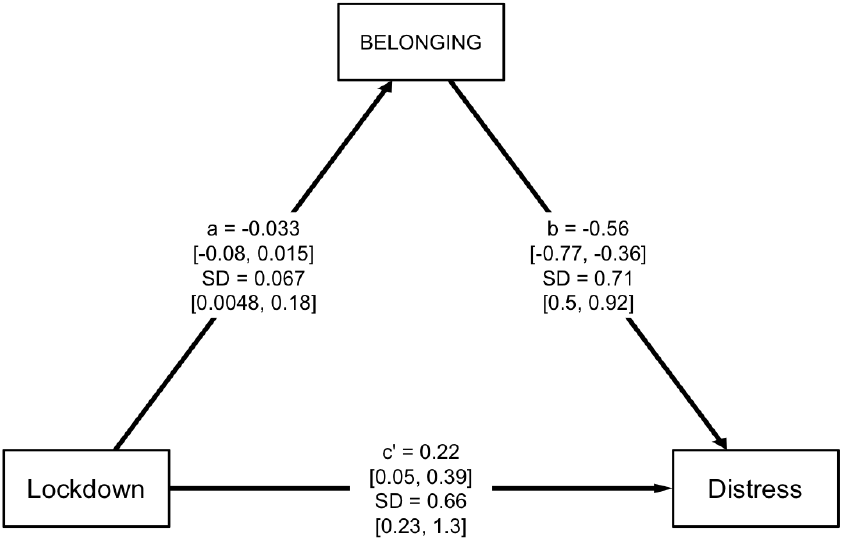
At the population-level average, the lockdown did not reliably affect perceived social support; had it done so, psychological distress would have been worse.

**Figure F10.**
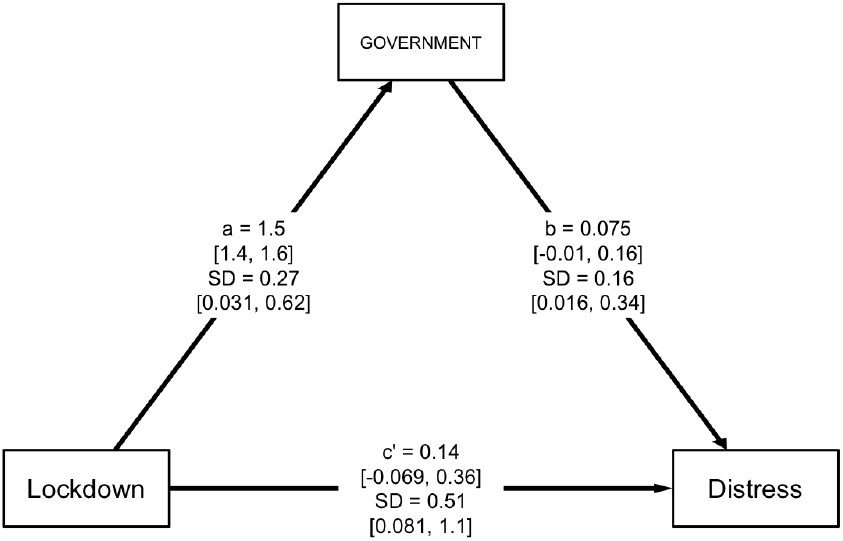
At the population-level average, the lockdown did not reliably affect perceived social belonging; had it done so, psychological distress would have been worse.

**Figure F11.**
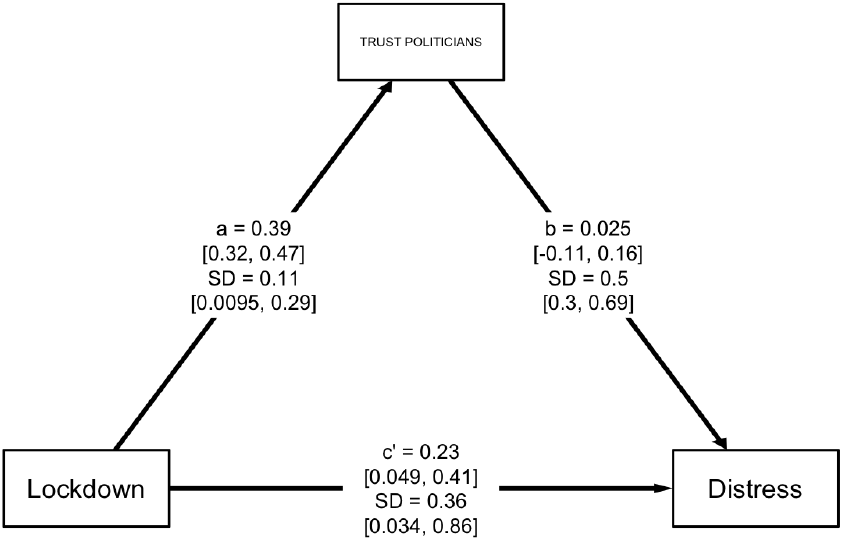
At the population-level average, the lockdown reduced satisfaction with personal relationships; moreover, dissatisfaction with personal relationships resulting from lockdown fully mediated observed psychological distress.

**Figure F12.**
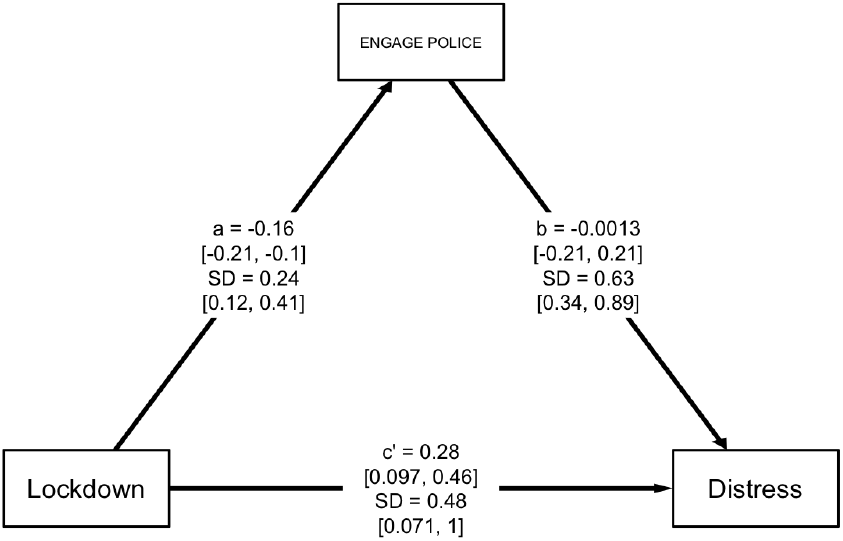
At the population-level average, the lockdown strongly in-creased satisfaction with the performance of government; however, satisfaction with government did not reliably predict psychological distress.

**Figure F13.**
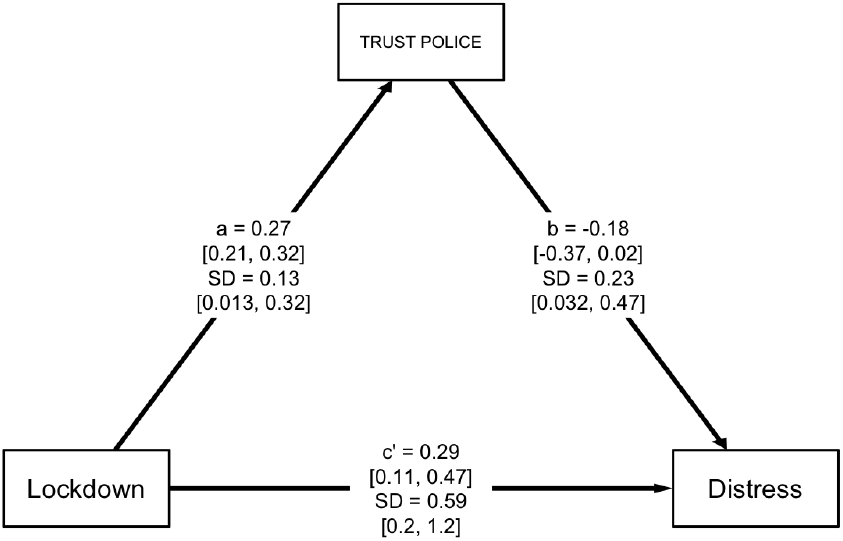
At the population-level average, the lockdown increased trust in politicians; however, trust in politicians did not reliably predict psychological distress.

**Figure F14.**
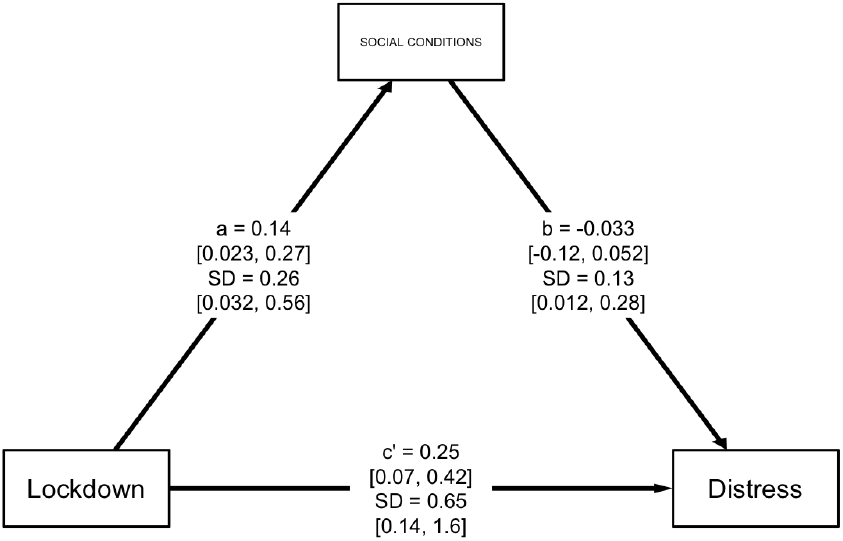
At the population-level average, the lockdown decreased willingness to engage with the police; however, willingness to engage with the police did not reliably predict psychological distress.

**Figure F15.**
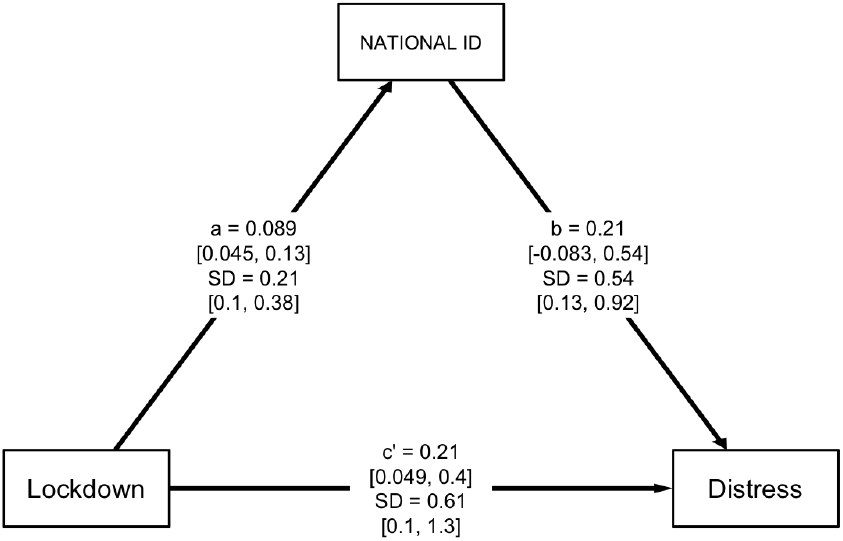
At the population-level average, the lockdown increased trust in the police; however, trust in the police did not reliably predict psychological distress.

**Figure F16.**
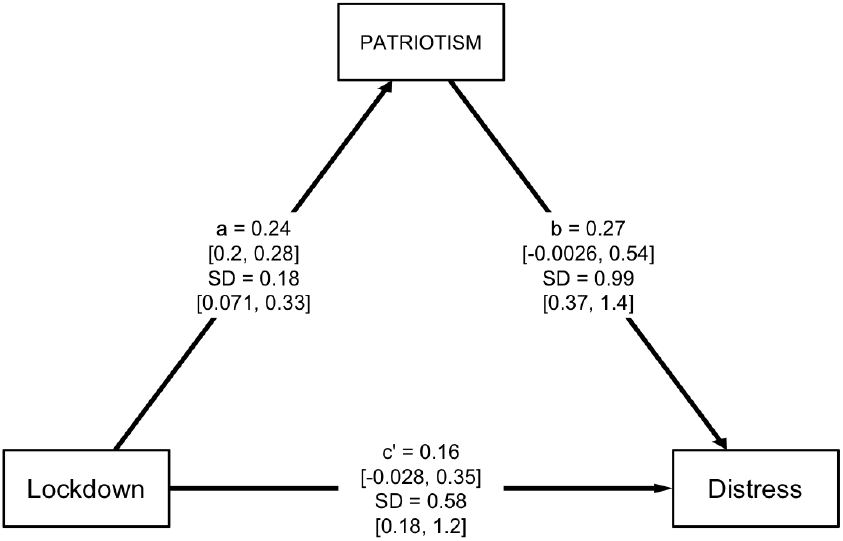
At the population-level average, the lockdown reliably increased satisfaction with social conditions; however, satisfaction with social conditions did not reliably predict psychological distress.

**Figure F17.**
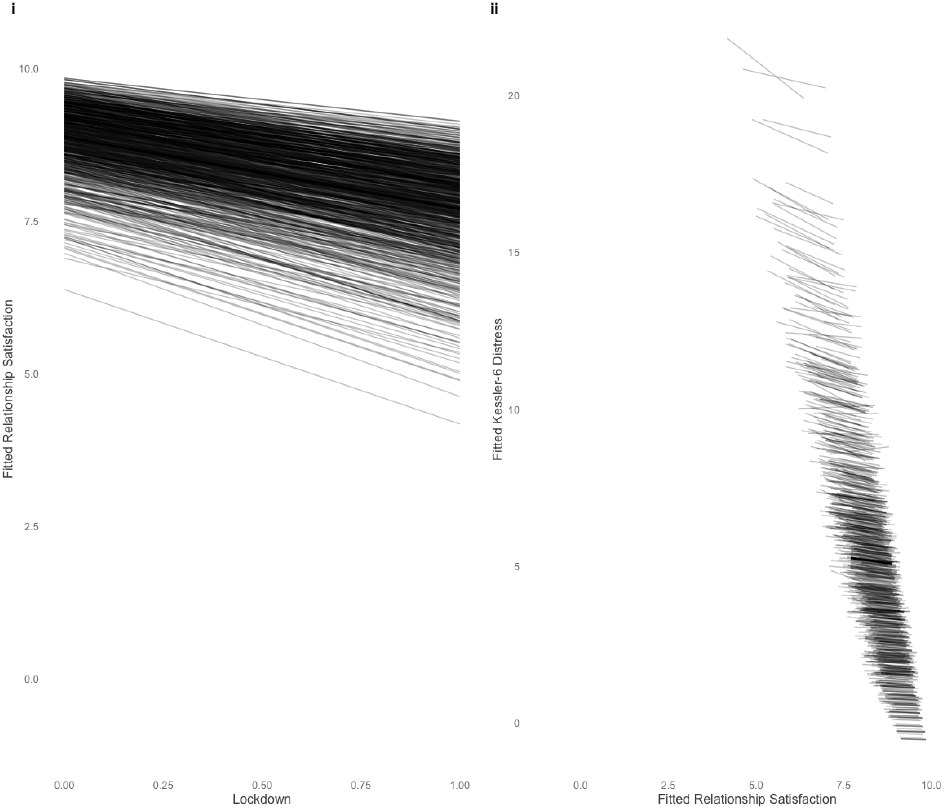
At the population-level average, the lockdown reliably increased feelings of national identity; however, national identity did not reliably predict distress.

**Figure F18.**
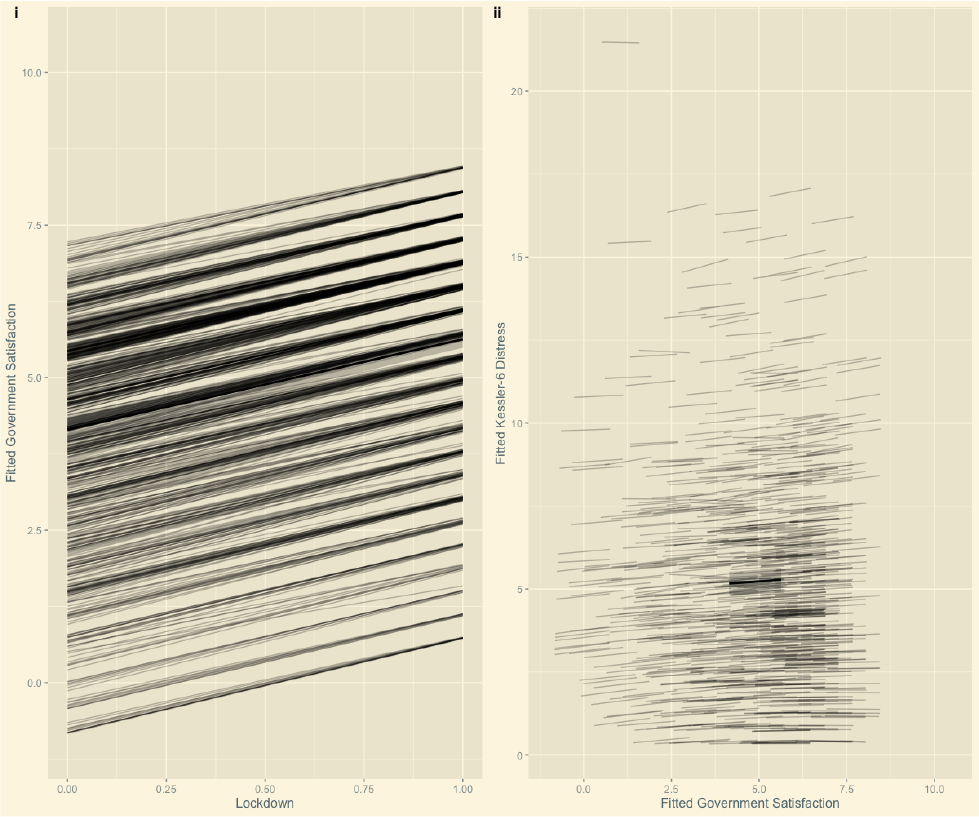
At the population-level average, the lockdown substantially increased patriotism; however, patriotism did not reliably predict distress.

**Figure F19.**
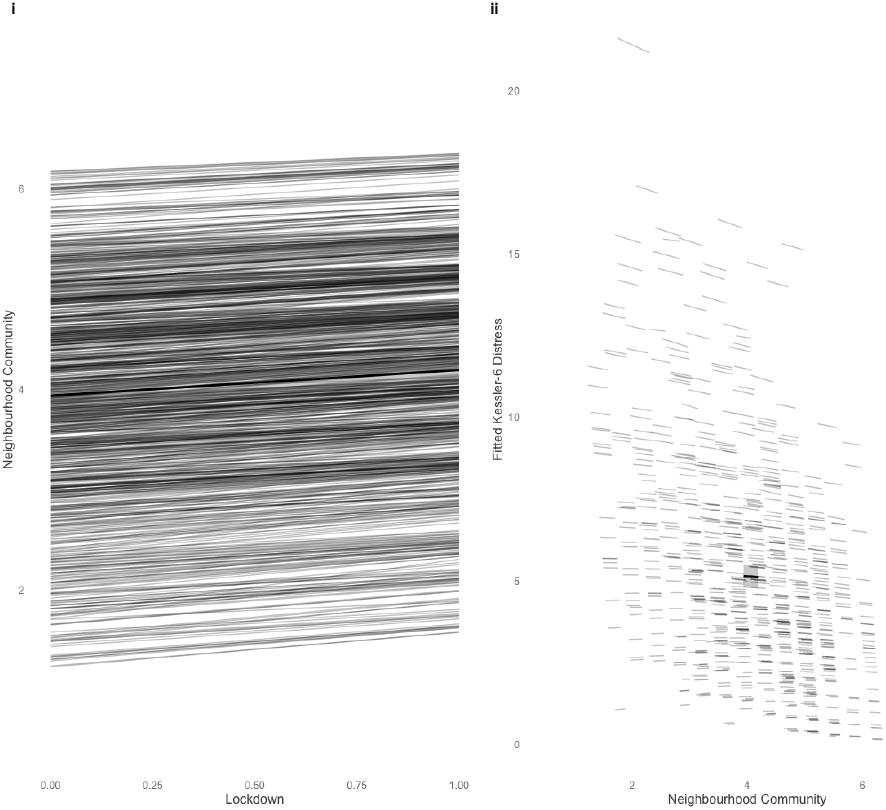
At the population-level average, the lockdown substantially increased a sense of neighbourhood community; moreover, a sense of neighbourhood community reliably buffered people from greater psychological distress.

## Appendix G Results in Tables

**Table G1.** Health Satisfaction

| Parameter | Mean | 5% | 95% |
| --- | --- | --- | --- |
| a | -0.07 | -0.17 | 0.04 |
| b | -0.16 | -0.26 | -0.06 |
| cp | 0.24 | 0.07 | 0.41 |
| me | 0.01 | -0.03 | 0.04 |
| c | 0.25 | 0.07 | 0.42 |
| tau_a | 0.15 | 0.01 | 0.42 |
| tau_b | 0.27 | 0.17 | 0.38 |
| tau_cp | 0.75 | 0.28 | 1.45 |
| corrab | -0.04 | -0.65 | 0.61 |

**Table G2.** Short Form Health

| Parameter | Mean | 5% | 95% |
| --- | --- | --- | --- |
| a | 0.04 | -0.02 | 0.09 |
| b | -0.48 | -0.68 | -0.28 |
| cp | 0.26 | 0.09 | 0.43 |
| me | -0.02 | -0.06 | 0.03 |
| c | 0.24 | 0.07 | 0.41 |
| tau_a | 0.07 | 0.00 | 0.21 |
| tau_b | 0.69 | 0.45 | 0.92 |
| tau_cp | 0.51 | 0.13 | 1.07 |
| corrab | 0.03 | -0.62 | 0.66 |

**Table G3.** Rumination

| Parameter | Mean | 5% | 95% |
| --- | --- | --- | --- |
| a | -0.03 | -0.08 | 0.02 |
| b | 1.53 | 1.33 | 1.72 |
| cp | 0.32 | 0.17 | 0.47 |
| me | -0.06 | -0.16 | 0.03 |
| c | 0.26 | 0.09 | 0.42 |
| tau_a | 0.12 | 0.02 | 0.27 |
| tau_b | 1.00 | 0.81 | 1.19 |
| tau_cp | 0.51 | 0.08 | 0.95 |
| corrab | -0.16 | -0.68 | 0.38 |

**Table G4.** Fatigue

| Parameter | Mean | 5% | 95% |
| --- | --- | --- | --- |
| a | -0.21 | -0.26 | -0.16 |
| b | 0.90 | 0.72 | 1.09 |
| cp | 0.44 | 0.27 | 0.60 |
| me | -0.19 | -0.27 | -0.11 |
| c | 0.25 | 0.08 | 0.42 |
| tau_a | 0.11 | 0.01 | 0.29 |
| tau_b | 0.93 | 0.77 | 1.09 |
| tau_cp | 0.22 | 0.02 | 0.57 |
| corrab | -0.01 | -0.61 | 0.54 |

**Table G5.** Satisfied with the Economy

| Parameter | Mean | 5% | 95% |
| --- | --- | --- | --- |
| a | 0.06 | -0.07 | 0.19 |
| b | -0.05 | -0.13 | 0.02 |
| cp | 0.23 | 0.06 | 0.40 |
| me | 0.01 | -0.02 | 0.05 |
| c | 0.24 | 0.07 | 0.41 |
| tau_a | 0.24 | 0.03 | 0.54 |
| tau_b | 0.19 | 0.08 | 0.31 |
| tau_cp | 0.52 | 0.11 | 1.10 |
| corrab | 0.21 | -0.44 | 0.77 |

**Table G6.**
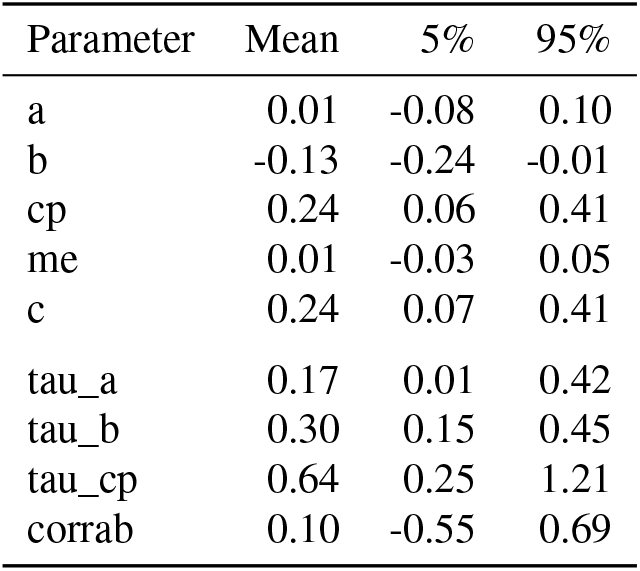
Satisfied with Standard of Living

| Parameter | Mean | 5% | 95% |
| --- | --- | --- | --- |
| a | 0.01 | -0.08 | 0.10 |
| b | -0.13 | -0.24 | -0.01 |
| cp | 0.24 | 0.06 | 0.41 |
| me | 0.01 | -0.03 | 0.05 |
| c | 0.24 | 0.07 | 0.41 |
| tau_a | 0.17 | 0.01 | 0.42 |
| tau_b | 0.30 | 0.15 | 0.45 |
| tau_cp | 0.64 | 0.25 | 1.21 |
| corrab | 0.10 | -0.55 | 0.69 |

**Table G7.** Satisfied with Future Security

| Parameter | Mean | 5% | 95% |
| --- | --- | --- | --- |
| a | -0.10 | -0.22 | 0.03 |
| b | -0.11 | -0.20 | -0.02 |
| cp | 0.24 | 0.06 | 0.42 |
| me | 0.01 | -0.06 | 0.08 |
| c | 0.25 | 0.07 | 0.42 |
| tau_a | 0.32 | 0.07 | 0.61 |
| tau_b | 0.40 | 0.28 | 0.51 |
| tau_cp | 0.54 | 0.13 | 1.11 |
| corrab | -0.01 | -0.56 | 0.54 |

**Table G8.** Satisfied with Business

| Parameter | Mean | 5% | 95% |
| --- | --- | --- | --- |
| a | -0.32 | -0.44 | -0.20 |
| b | -0.04 | -0.14 | 0.05 |
| cp | 0.25 | 0.07 | 0.44 |
| me | -0.01 | -0.07 | 0.06 |
| c | 0.24 | 0.07 | 0.42 |
| tau_a | 0.69 | 0.50 | 0.93 |
| tau_b | 0.16 | 0.02 | 0.31 |
| tau_cp | 0.73 | 0.29 | 1.41 |
| corrab | -0.19 | -0.68 | 0.36 |

**Table G9.** Social Support

| Parameter | Mean | 5% | 95% |
| --- | --- | --- | --- |
| a | 0.03 | -0.03 | 0.08 |
| b | -0.49 | -0.70 | -0.27 |
| cp | 0.24 | 0.07 | 0.41 |
| me | 0.00 | -0.05 | 0.07 |
| c | 0.24 | 0.07 | 0.41 |
| tau_a | 0.10 | 0.01 | 0.26 |
| tau_b | 0.76 | 0.48 | 1.03 |
| tau_cp | 0.52 | 0.09 | 1.15 |
| corrab | 0.10 | -0.55 | 0.67 |

**Table G10.** Social Belonging

| Parameter | Mean | 5% | 95% |
| --- | --- | --- | --- |
| a | -0.03 | -0.08 | 0.01 |
| b | -0.56 | -0.77 | -0.36 |
| cp | 0.22 | 0.05 | 0.39 |
| me | 0.02 | -0.02 | 0.07 |
| c | 0.24 | 0.07 | 0.41 |
| tau_a | 0.07 | 0.00 | 0.18 |
| tau_b | 0.71 | 0.50 | 0.92 |
| tau_cp | 0.66 | 0.23 | 1.30 |
| corrab | 0.08 | -0.57 | 0.68 |

**Table G11.** Satisfied with Personal Relationships

| Parameter | Mean | 5% | 95% |
| --- | --- | --- | --- |
| a | -1.18 | -1.33 | -1.03 |
| b | -0.16 | -0.24 | -0.07 |
| cp | 0.05 | -0.15 | 0.24 |
| me | 0.20 | 0.09 | 0.32 |
| c | 0.25 | 0.08 | 0.42 |
| tau_a | 0.61 | 0.34 | 0.89 |
| tau_b | 0.27 | 0.16 | 0.38 |
| tau_cp | 1.30 | 0.75 | 2.15 |
| corrab | 0.12 | -0.32 | 0.55 |

**Table G12.** Satisfied with the Performance of Government

| Parameter | Mean | 5% | 95% |
| --- | --- | --- | --- |
| a | 1.48 | 1.36 | 1.60 |
| b | 0.07 | -0.01 | 0.16 |
| cp | 0.14 | -0.07 | 0.36 |
| me | 0.10 | -0.03 | 0.23 |
| c | 0.25 | 0.08 | 0.42 |
| tau_a | 0.27 | 0.03 | 0.62 |
| tau_b | 0.16 | 0.02 | 0.34 |
| tau_cp | 0.51 | 0.08 | 1.08 |
| corrab | -0.12 | -0.73 | 0.55 |

**Table G13.** Trust Politicians

| Parameter | Mean | 5% | 95% |
| --- | --- | --- | --- |
| a | 0.39 | 0.32 | 0.47 |
| b | 0.02 | -0.11 | 0.16 |
| cp | 0.23 | 0.05 | 0.41 |
| me | 0.01 | -0.05 | 0.08 |
| c | 0.24 | 0.07 | 0.42 |
| tau_a | 0.11 | 0.01 | 0.29 |
| tau_b | 0.50 | 0.30 | 0.69 |
| tau_cp | 0.36 | 0.03 | 0.86 |
| corrab | 0.01 | -0.64 | 0.64 |

**Table G14.** Willingness to Engage with Police

| Parameter | Mean | 5% | 95% |
| --- | --- | --- | --- |
| a | -0.16 | -0.21 | -0.10 |
| b | 0.00 | -0.21 | 0.21 |
| cp | 0.28 | 0.10 | 0.46 |
| me | -0.03 | -0.10 | 0.04 |
| c | 0.25 | 0.08 | 0.42 |
| tau_a | 0.24 | 0.12 | 0.41 |
| tau_b | 0.63 | 0.34 | 0.89 |
| tau_cp | 0.48 | 0.07 | 1.01 |
| corrab | -0.18 | -0.60 | 0.30 |

**Table G15.** Trust Police

| Parameter | Mean | 5% | 95% |
| --- | --- | --- | --- |
| a | 0.27 | 0.21 | 0.32 |
| b | -0.18 | -0.37 | 0.02 |
| cp | 0.29 | 0.11 | 0.47 |
| me | -0.05 | -0.11 | 0.01 |
| c | 0.24 | 0.07 | 0.42 |
| tau_a | 0.13 | 0.01 | 0.32 |
| tau_b | 0.23 | 0.03 | 0.47 |
| tau_cp | 0.59 | 0.20 | 1.16 |
| corrab | 0.00 | -0.65 | 0.64 |

**Table G16.** Satisfied with Social Conditions

| Parameter | Mean | 5% | 95% |
| --- | --- | --- | --- |
| a | 0.14 | 0.02 | 0.27 |
| b | -0.03 | -0.12 | 0.05 |
| cp | 0.25 | 0.07 | 0.42 |
| me | 0.00 | -0.03 | 0.04 |
| c | 0.25 | 0.07 | 0.42 |
| tau_a | 0.26 | 0.03 | 0.56 |
| tau_b | 0.13 | 0.01 | 0.28 |
| tau_cp | 0.65 | 0.14 | 1.63 |
| corrab | 0.14 | -0.55 | 0.74 |

**Table G17.**
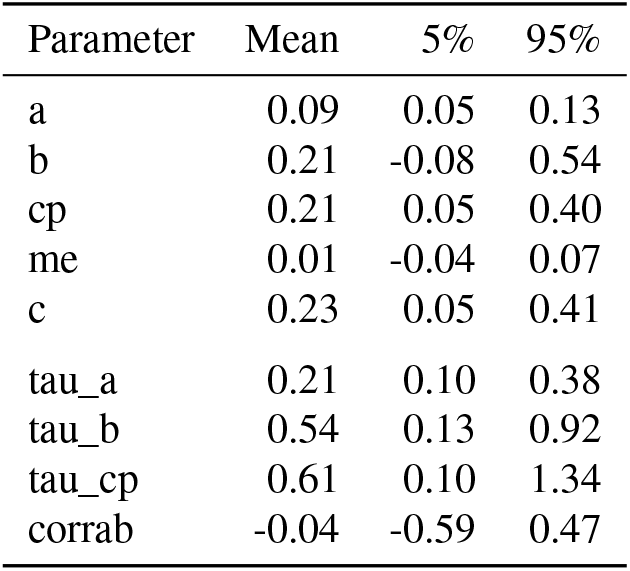
National Identity

**Table G18.** Patriotism

| Parameter | Mean | 5% | 95% |
| --- | --- | --- | --- |
| a | 0.24 | 0.20 | 0.28 |
| b | 0.27 | 0.00 | 0.54 |
| cp | 0.16 | -0.03 | 0.35 |
| me | 0.08 | -0.01 | 0.19 |
| c | 0.24 | 0.07 | 0.41 |
| tau_a | 0.18 | 0.07 | 0.33 |
| tau_b | 0.99 | 0.37 | 1.36 |
| tau_cp | 0.58 | 0.18 | 1.19 |
| corrab | 0.11 | -0.37 | 0.56 |

**Table G19.** Sense of Neighbourhood Community

| Parameter | Mean | 5% | 95% |
| --- | --- | --- | --- |
| a | 0.25 | 0.18 | 0.33 |
| b | -0.13 | -0.26 | -0.01 |
| cp | 0.30 | 0.12 | 0.48 |
| me | -0.05 | -0.11 | 0.00 |
| c | 0.25 | 0.07 | 0.43 |
| tau_a | 0.18 | 0.02 | 0.43 |
| tau_b | 0.28 | 0.12 | 0.45 |
| tau_cp | 0.40 | 0.05 | 0.92 |
| corrab | -0.26 | -0.79 | 0.41 |

